# Genome-wide profiling of short tandem repeat somatic instability reveals associations with age, sex and brain-related traits

**DOI:** 10.1101/2025.11.13.25340159

**Authors:** Jialu Hu, Richard Mantey, Hengyuan Guo, Ahmed M. Sidky, Maryam Touhidinia, Tanzeem Butt, Roohollah Sobhani, Santiago Estrada, Kristian Haendler, Shixiao Zhao, Xuequn Shang, Elena De Domenico, Marc D. Beyer, Monique M. B. Breteler, N. Ahmad Aziz

**Author notes:** **Corresponding author:** Prof. Dr. N. Ahmad Aziz, MD, PhD Population & Clinical Neuroepidemiology, German Center for Neurodegenerative Diseases (DZNE) Venusberg-Campus 1/99, Bonn 53127, Germany.

## Abstract

Short tandem repeats (STRs) are highly abundant in the human genome and their age-related somatic instability is emerging as a pivotal pathomechanism in repeat expansion disorders. Nevertheless, currently no analysis tool exists for genome-wide profiling of STR somatic instability. Here, we present *searchSTR*, a novel computational framework that enables accurate STR genotyping and somatic instability quantification from both whole-genome and targeted sequencing data. Applying this approach, we analyzed STR variants from whole-genome sequencing data of the 1000 Genomes Project (n=3,202) and targeted deep-sequencing data of the population-based Rhineland Study (n=2,974). We provide a comprehensive multiancestry genome-wide reference panel of STR variants and their somatic instability, covering more than 1.4 million STRs across 26 populations. Remarkably, STR somatic instability at many loci was robustly associated with age, sex, brain morphology and markers of neurodegeneration. These findings reveal a crucial role for STR somatic instability as an age-dependent modifier of brain structure.

## Introduction

Short tandem repeats (STRs), also referred to as microsatellites, are abundant, highly polymorphic DNA sequences that consist of tandemly repeated 1–6 bp motifs [1]. When the repeating unit extends beyond 6 bp, typically ranging from 7–100 bp, these sequences are commonly referred to as variable number tandem repeats (VNTRs) or macrosatellites. Together, tandem repeats represent one of the most prevalent classes of genetic variation in humans, with STRs alone constituting an estimated 6.77% of the genome, spanning more than 1.5 million annotated loci [1, 2]. STR variation encompasses changes not only in the number of repeat units but also in the motif structure and underlying sequence composition. Such polymorphisms exert profound functional effects, ranging from subtle alterations in gene expression to catastrophic disruptions of genome integrity [3]. The mutability of STRs, which can be several orders of magnitude higher than that of single-nucleotide variants, renders them a major source of both evolutionary adaptation and pathogenic instability [4]. Clinically, STR expansions are known to cause a broad class of diseases referred to as repeat expansion disorders (REDs) [5]. To date, repeat expansions have been implicated in the pathogenesis of at least 65 neurological and 14 neuromuscular disorders, underscoring their extraordinary clinical relevance [6]. For example, trinucleotide expansions of CAG repeats within coding regions of several different genes result in an elongated polyglutamine (polyQ) tract in the resultant protein products, and account for a family of nine polyQ diseases [7], including Huntington’s disease (HD), spinocerebellar ataxias (SCA1, SCA2, SCA3, SCA6, SCA7, and SCA17), dentatorubral-pallidoluysian atrophy (DRPLA) and spinal-bulbar muscular atrophy (SBMA). Similarly, pathological expansions of CCG repeats in the *FMR1* gene lead to fragile X syndrome [4], the most common genetic cause of intellectual disability. In addition, STR size variations can act as modifiers of disease expression in a range of neuromuscular and neuropsychiatric disorders such as amyotrophic lateral sclerosis and autism spectrum disorders [8].

Apart from germline STR variations, emerging evidence indicates that also somatic STR expansions play a critical role in modifying disease phenotype. For example, in a landmark study it was demonstrated that increases in somatic CAG repeat expansion ratio over time in blood were a significant predictor of subsequent caudate and putamen atrophy in HD mutation carriers [9]. Unlike germline STR variations, which are inherited and broadly consistent across tissues, somatic expansions occur post-zygotically and can vary widely across cell types and even within specific cell populations from the same tissue [10]. This mosaicism introduces significant variability in disease phenotype, influencing the onset, progression, and severity of several REDs [11]. Moreover, somatic STR expansions often exhibit age-dependent accumulation [12, 13], correlating with disease progression and providing a mechanistic link between genetic instability and clinical deterioration. Understanding the determinants of somatic STR expansions is crucial, as they not only impact the molecular pathways driving disease, but also hold great potential as biomarkers for disease staging and targets for therapeutic intervention. The highly dynamic nature of STR expansions highlights the necessity of developing advanced computational algorithms capable of capturing their complexity. Such approaches should account for tissue-specific variability and longitudinal changes across the lifespan, enabling a more accurate characterization of expansion patterns. Ultimately, these methodological advances will provide critical insights into disease mechanisms and open avenues for more precise, personalized strategies for both diagnosis and therapy.

To date, most computational methods for STR analysis have concentrated on detecting and genotyping germline STR variations in population and evolutionary studies. Consequently, systematic approaches for characterizing somatic STR expansion are still lacking. Early algorithms developed for STR genotyping, such as LobSTR [14] and RepeatSeq [15], require that target STR alleles be contained within the read length. Although they produce high-confidence genotypes, they are unable to call STR alleles beyond the read length, including most pathogenic expansions. With the advent of long-read whole-genome sequencing (WGS) technologies (e.g., PacBio Single Molecule Real-Time sequencing and Oxford Nanopore Technology), long-read methods have expanded the detection of variants in longer tandem repeats [16, 17], including VNTRs, and can be used to assemble multi-megabase satellite repeats. However, long-read sequencing is limited by lower throughput, higher per-base error rate and higher costs than short-read sequencing. TREDPARSE [18] incorporates various cues from read alignment and pair-end distance distribution to infer STR repeat size from short-read sequencing data, although it is limited to a small number of STR loci. To address the above limitations, two other STR genotyping tools, ExpansionHunter [19] and GangSTR [20] were developed to call moderately longer STR alleles from short-read WGS data. ExpansionHunter utilizes several probabilistic models to estimate repeat sizes by analysing three types of identified reads—spanning reads, flanking reads, and in-repeat reads—within the maximum likelihood estimation (MLE) framework. GangSTR estimates the repeat size based on a unified model within the MLE framework. More recently, STRling [21] was developed, which leverages the efficiency of *k*-mer counting to identify previously uncharacterized STR loci, enabling the discovery of new repeat expansions. However, because this method counts all substrings of a specified length, *k*-mers that originate from both the repeat and the flanking regions can be misclassified, leading to uncertainty in determining the true repeat boundaries. This limitation can make it difficult to resolve complex repeat structures or distinguish between similar repetitive regions in the genome, particularly in areas with high sequence homology or multiple nearby STR loci. Leveraging individual STR genotyping tools or a combination of them, several pioneering works [22–24] have extensively investigated genome-wide STR variation across several population-based and clinical cohorts, including TOPMed [25], UK Biobank, 1000 Genomes Project [26], NyuWa [27], Simons Simplex Collection [28], and All of US Research Program [29].

Recently, the development of therapeutic strategies targeting STR somatic instability has emerged as one of the most active and promising areas of research in REDs. However, despite the critical need for reliable high-throughput profiling of STR somatic instability from (short-read) WGS or targeted sequencing data, currently no computational frameworks are available for accomplishing this pivotal task. This limitation arises from several technical and methodological challenges. First, the detection of STR variations is inherently difficult due to low capture rate, allelic dropout in the extraction of low abundant DNA from samples, and sequencing technology errors. Second, the low sequence complexity nature of STRs and their flanking regions may lead to alignment ambiguities, and reduced efficiency in inferring diploid repeat lengths, requiring new genome-wide catalogues of STRs with high-complexity flanking regions for accurate mapping. Although single-cell genome sequencing data theoretically could provide a direct way to study mutations at single-cell resolution in germline and somatic tissues independently, single-cell genomic sequencing methods suffer from high rates of dropout and artefacts during whole-genome amplification [30]. Third, most STR genotyping algorithms estimate diploid repeat lengths using a mixture of symmetric probability models to fit the repeat counting data, which may lead to under- or overestimation due to the prevalent nature of unbalanced STR expansion and contraction rates. Due to these technical and methodological limitations, current understanding of the patterns of somatic STR mutations across cell types, and their impact on cell fates and phenotypes, remains limited. Thus, there is a pressing need for novel computational approaches capable of accurately identifying and characterizing somatic STR variations.

To address the critical gaps detailed above, we developed *searchSTR,* an integrated workflow that incorporates a suite of innovative methodological and algorithmic strategies, enabling comprehensive characterization of STR variations and their somatic instability from either WGS or targeted sequencing data. This included the development of a novel STR genotyping tool that can call somatic STR variations from genome sequencing data of individuals based on the input of a tailored STR catalogue, enabling haplotype-specific STR length estimates. Unlike other existing tools that use symmetric probability models for inference of diploid repeat lengths, we employed a mixture of two skewed distributions within a MLE framework to accurately infer the most likely repeat units for each of the germline alleles. Taking advantage of the fast algorithmic component in the SeqAn library [31], our software is computationally efficient and can be deployed in large population-scale cohorts. Indeed, application of our method to the WGS data of the 1000 Genomes Project yielded a comprehensive multiancestry genome-wide reference panel of STR somatic instability. Moreover, using targeted deep-sequencing data from the population-based Rhineland Study, we found STR somatic instability at many loci to be robustly associated with age, sex, brain morphology and markers of neurodegeneration, demonstrating the important but hitherto unappreciated role of STR somatic instability as an age-dependent modifier of brain structure.

## Results

### Overview of searchSTR/STRbean

We developed *searchSTR*, an open-source, ultrafast, and haplotype-resolved method implemented in the *STRbean* package [32], for accurate detection, comprehensive analysis, and functional characterization of genome-wide STR variants and their somatic instability from short-read sequencing data at population scale (a schematic overview of our approach is presented in *Fig. 1A*). Our tool implements thread-level parallelism and can be further accelerated by incorporating chromosome-level parallelism. Our method incorporates a filter-based pipeline, bucket hashing and mixture models within the MLE framework to estimate allele-specific STR length and somatic instability at repeat unit length for each individual. We applied *searchSTR* to WGS data from 3,202 samples in the 1000 Genomes Project to characterize over 1.6 million STR variants, as well as to targeted deep-sequencing data on a curated panel of 2,749 genomic regions – selected for clinical and functional relevance – in 2,974 participants of the population-based Rhineland Study. Leveraging the deep-phenotyping background data from the Rhineland Study, we also assessed the association between somatic expansion instability (SEI) of 4,823 STR variants and 11 other traits (including age, sex, eight brain imaging-derived phenotypes as well as plasma levels of neurofilament light chain (NfL), an ultrasensitive marker of neurodegeneration) using multiple linear regression and Bayesian zero-inflated Gaussian models. Apart from a range of technical improvements for accurate calling of STR size and identifying pathogenic STR expansions, the major advantage of our method is that in contrast to all existing STR genotyping tools, it provides genome-wide estimates of somatic instability of STR loci. Source code of our package is freely available at https://github.com/jhu99/STRbean under BSD 3-clause license.

**Figure 1.**
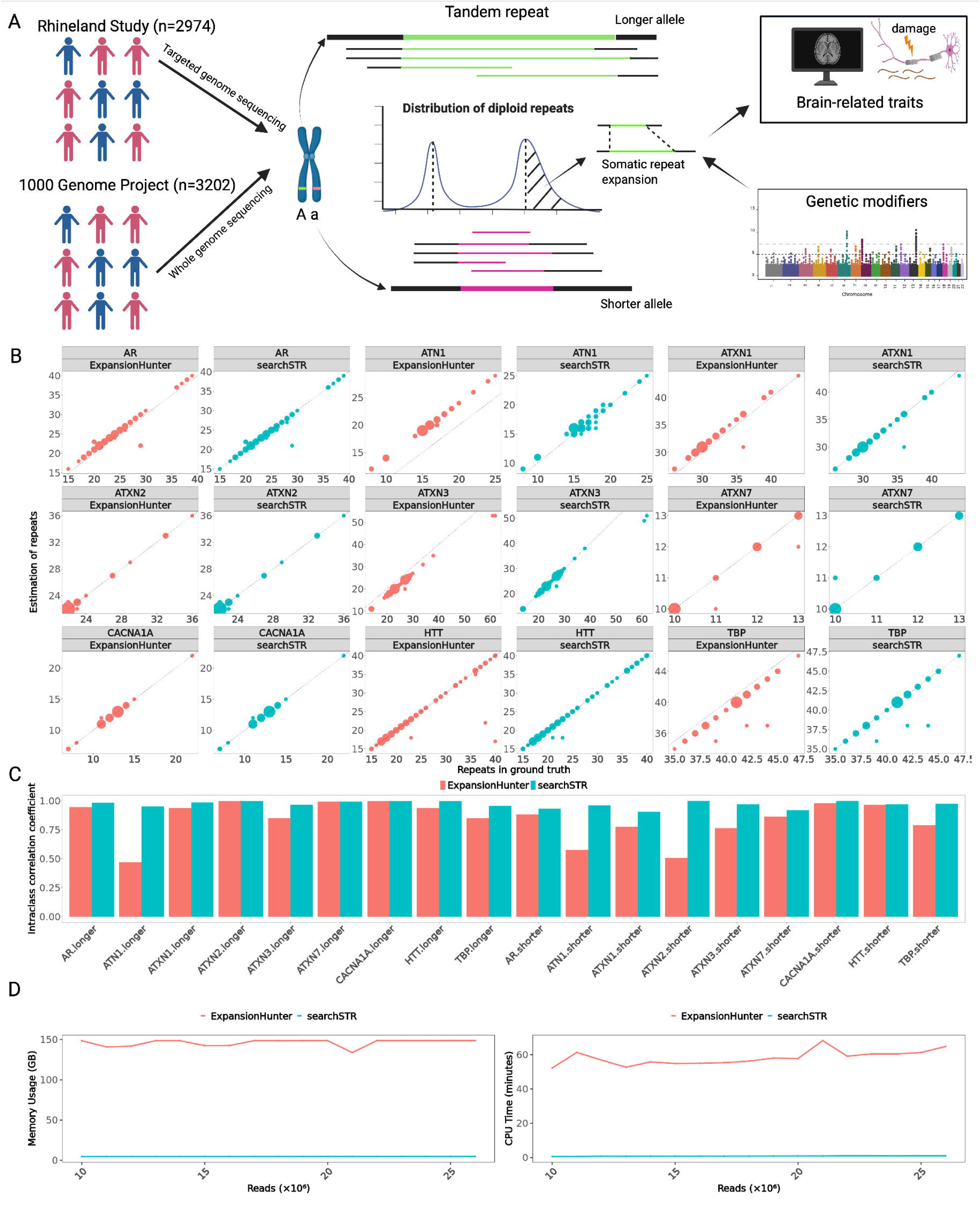
Workflow and comparison of searchSTR with the current state-of-the-art for STR repeat size estimation. (A) Schematic overview of STR genotyping and association analyses between somatic expansion and brain-related traits. (B) Benchmark analysis of STR length estimation for the longer allele at nine polyglutamine disease–associated loci using searchSTR (cyan) and ExpansionHunter (red). The x-axis represents the ground truth (STR size measured by PCR-based fragment analysis). Each dot denotes a group of individuals with the same repeat size, with dot size indicating the number of samples in that group. (C) Performance comparison based on agreement between estimated repeat lengths and PCR-based fragment analysis, assessed using intraclass correlation coefficients (ICC). (D) Comparison of searchSTR and ExpansionHunter in terms of computational performance. Memory usage (left) and CPU time (right) were measured across datasets with increasing sequencing read counts.

### Targeted genome sequencing data

To assess its performance on panel-based data, we applied *searchSTR* to targeted deep-sequencing data from 2,974 participants of the Rhineland Study. Genomic DNA was extracted from peripheral whole blood samples, and sequencing was performed using an amplicon-based targeted approach on the Illumina NovaSeq 6000 platform. The panel targeted 2,749 genomic regions of GRCh38 (*Supplementary Tab. 1*), including most of the known RED-causing loci, as well as additional loci selected based on their potential associations with brain function and neurodegenerative disease from prior comparative genomic analyses [33–35]. Highly polymorphic microsatellite regions commonly used in forensic genotyping and DNA fingerprinting were also included to maximize allelic diversity in the target space. The sequencing protocol generated four paired-end FASTQ files per individual (two lanes × forward/reverse reads, 2×250 bp), providing high-quality sequencing data suitable for STR genotyping.

### Assessment of diploid STR detection accuracy across benchmark datasets using *searchSTR* and *ExpansionHunter*

To assess the accuracy of *searchSTR* in predicting diploid STR allele lengths, we compared it against the current state-of-the-art method, *ExpansionHunter* (v5.0.0), to determine repeat lengths at nine polyQ disease-associated STR loci in *AR, ATN1, ATXN1, ATXN2, ATXN3, ATXN7, CACNA1A, HTT* and *TBP* genes, from targeted genome sequencing data derived from ∼170 individuals of two well-characterized Dutch cohorts [36]. The genomic coordinates corresponding to the nine STR loci were obtained from the Illumina Repeat Catalogs [37]. The selected loci represent well-characterized pathogenic STRs and serve as a standardized benchmark for evaluating STR genotyping tools. A PCR-based fragment analysis served as the ground truth for determining repeat lengths as detailed previously [38]. The results in *Fig. 1C* demonstrate that *searchSTR* is robust in estimating repeat lengths across both the longer (*Fig. 1B*) and shorter (*Supplementary Fig. 1*) alleles of these nine STR loci. Our benchmark analyses further revealed that *searchSTR* outperforms *ExpansionHunter* in terms of intraclass correlation coefficients (ICCs) with the ground truth, indicating superior reliability and consistency in estimating the repeat lengths for both alleles across the nine polyQ disease-associated loci. Notably, the accuracy of *searchSTR* substantially exceeded that of *ExpansionHunter* for the longer allele of *ATN1* as well as for the shorter alleles of *ATN1*, *ATXN2*, *ATXN3*, and *TBP*. This improvement likely results from the adoption of our new non-symmetric model (*Equations 1,2*). Analyses of sequencing read distributions from these nine polyQ disease–associated loci in two Dutch cohorts demonstrated that the frequencies of contracted and expanded reads are intrinsically non-symmetric. At five loci (*HTT*, *TBP*, *ATXN1*, *ATXN2*, and *ATXN3*), the distributions were consistently left-skewed, whereas at two loci (*ATN1* and *ATXN7*), they were right-skewed. These results highlight the utility of *searchSTR* for precise and reliable diploid STR genotyping, particularly in the context of pathogenic repeat expansions relevant to neurodegenerative diseases. In additional benchmarking tests, *searchSTR* completed the analysis of ∼20 million reads for a catalog containing 1.6 million STR loci in less than one minute with 2 threads, consuming approximately 5 GB of memory. As shown in *Fig. 1D*, on the same platform (Intel(R) Xeon(R) CPU E5-2630 v4) and across datasets with increasing reads (10-26 millions), *searchSTR* outperformed *ExpansionHunter* by up to two orders of magnitude in both speed and memory efficiency.

### Genome-wide catalogs of STR variation on GRCh38 and annotations

There are a dozen of general or disease-associated STR catalogs [22, 39, 40], which differ from one another because of variations in boundaries (without flanking regions), tolerance to mismatch (allowing repeat interruption), their definition of repeat patterns (simple motif or regular expression motif), reference genomes (hg19/hg38) and the focus on different tandem repeat classes (STR or VNTR). To generate a general whole-genome STR catalog that can meet different purposes, we developed a tool, called *STRfinder/STRbean* [32] that can facilitate generating a comprehensive catalog of STR variants for a reference genome based on custom designed tandem repeat definitions. Leveraging this tool, we generated two sets of genome-wide STR catalogs (i.e., exact and approximate catalogs) [41, 42] with different parameter settings on the human reference genome assembly build GRCh38. In brief, we successfully identified 1,656,159 STR intervals for the approximate catalog – including 589,924 (35.62%) homopolymers – of which 1,555,790 (93.94%) were located on autosomes, 82,736 (5.00%) on the X-chromosome, and 17,633 (1.06%) on the Y-chromosome, covering 34.2 Mbp (1.11%) of GRCh38. Similarly, 1,233,959 STR intervals were detected for the exact catalog – including 677,567 (54.91%) homopolymers – of which 1,162,690 (94.22%) were located on autosomes, 61,200 (4.96%) on the X-chromosome, and 10,069 (0.82%) on the Y-chromosome, covering 0.78% (24.2 Mbp) of GRCh38 (*Supplementary Tab. 2*). The STR density of both catalogs is visualized in *Fig. 2A,B*. Intervals that overlapped with known genes were automatically annotated with *STRfinder* based on gencode V38 [43]. A total of 940,059 (56.8%) STR loci from the approximate catalog were completely harbored within genes, including 21,771 STR loci within exons, 2,403 within 5’-UTRs, and 14,939 within 3’-UTRs. Similarly, 710,053 STR loci (57.5%) from the exact catalog were completely harbored within genes, including 14,348 within exons, 1,415 within 5’-UTRs, and 11,174 within 3’-UTRs (*Fig. 2C-E*). More details regarding the specific settings of *STRfinder* are presented in *Online Methods*.

**Figure 2.**
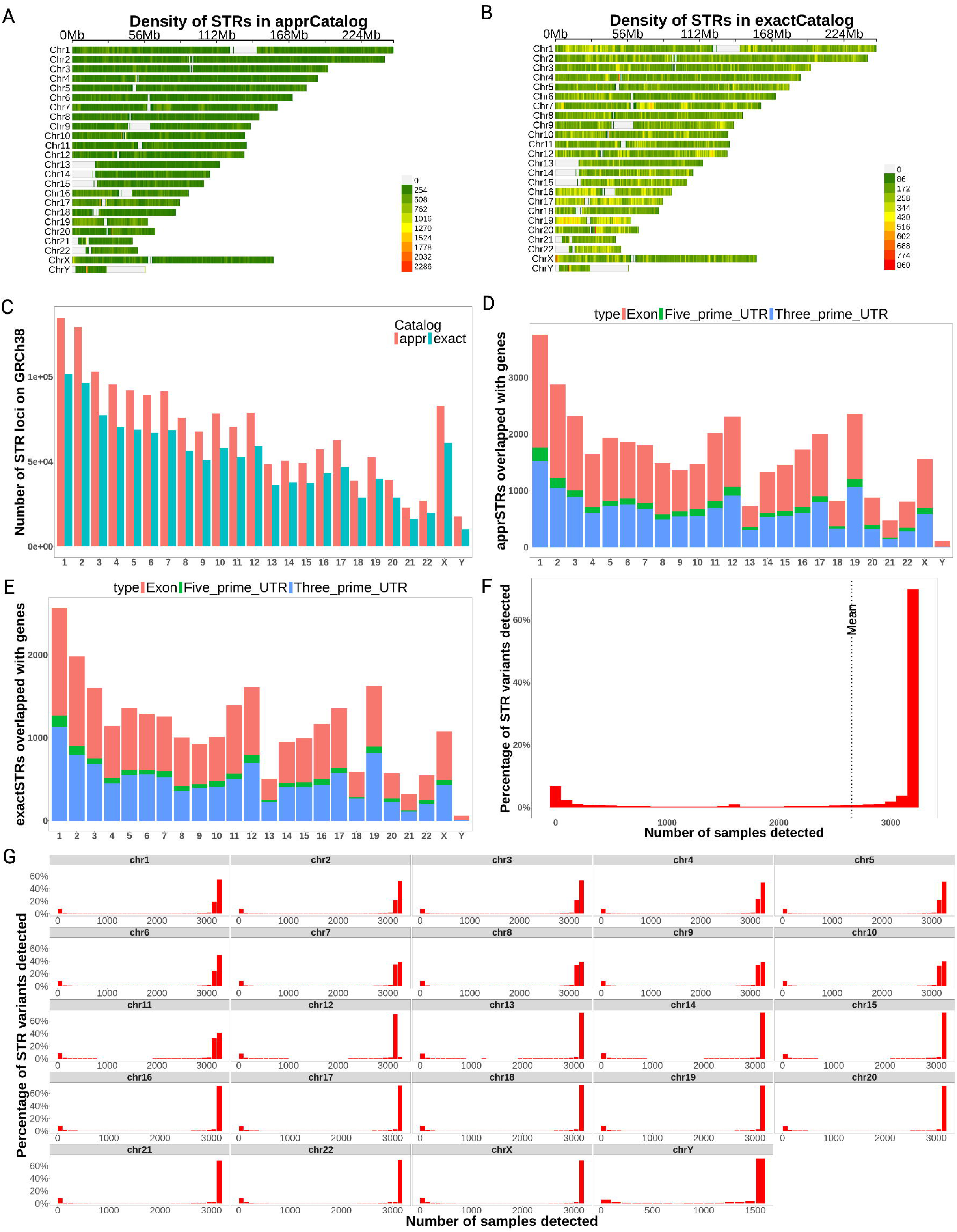
STR catalogs in GRCh38 and variants distribution in the 1000 Genome Project. (A) Chromosomal density of STR loci in apprCatalog. (B) Chromosomal density of STR loci in exactCatalog. (C) STR loci per chromosome in both catalogs (approximate repeats, red; exact repeats, cyan). Overlap of STR loci with gene features in apprCatalog (D) and exactCatalog (E). (F) Distribution of genotyped STR variants across individuals from the 1000 Genomes Project. (G) Percentage of genotyped STR variants per chromosome across samples. (unit: 5×10^5^ bp)

### Genome-wide STR genotyping across 3202 samples from 1000 Genomes Project

We also ran *searchSTR* on high-coverage (∼30x) PCR-free WGS data of 2,504 original samples and 698 additional related samples from the 1000 Genomes Project phase4 cohort [26]. This dataset includes 602 trios, 1,603 females, and 1,599 males. Using the approximate catalog generated above, for each individual *searchSTR* takes an alignment (BAM) file as input, detects paired reads that can be accurately mapped to a potential reference STR locus, and infers diploid repeat lengths for each STR variant (*Online Methods, Equations 1,2*), including *stutter* repeats. Several quality measures (e.g., at least 5 detected spanning reads) were used to remove poor-quality STR calls. In addition, call rates of variants on the Y-chromosome were calculated based on the male samples only. This resulted in the identification of a total of 1,400,982, 75,609 and 16,479 STR variants on the autosomes, and X- and Y-chromosome, respectively, each observed in at least one of the 3,202 analyzed samples. Of these, 1,112,549 variants were present with a call rate above 95%, while 1,202,985, 1,250,248, and 1,292,572 variants were detected in more than 75%, 50%, and 25% of the samples, respectively (*Supplementary Tab. 3, Fig. 2F*). At locus level, 1,112,973 STR variants were present in over 3000 samples (>93%), 123,650 variants were detectable in less than 100 samples (<3.1%), while 11,762 Y-chromosome variants were present in over 1500 male samples (>93.8%, *Fig. 2G*). In addition, *searchSTR* provides a comprehensive genotype call set for each STR locus across all analyzed individuals, including both allelic repeat lengths and the corresponding read support counts for each allele, reflecting the underlying sequencing evidence (*Supplementary Fig. 2A*).

Next, for each sample of the 2504 unrelated samples (female n=1271, male n=1233), we estimated a locus-specific SEI score to quantify allelic heterogeneity and mosaicism at each STR locus across the genome. *Fig. 3A, B, Suppl. Fig. 3A* show the average SEI Z-scores (avgSEI) across all loci, stratified by sex, five superpopulations and 26 populations. The results indicate that the AFR population exhibits the highest overall genomic instability, while males show higher SEI values than females. However, when restricting the avgSEI to autosomes, no significant sex differences were apparent (*Suppl. Fig. 2B*), indicating that the higher density of unstable STR expansions on the the Y chromosome underlies the overall higher somatic STR instability in males. A detailed description and definition of the SEI score is provided in *Online Methods (Equations 3)*. Furthermore, we characterized population-specific STR variants based on the longer of the two autosomal alleles at 1,400,982 loci across all superpopulations (AFR: African Ancestry n=661, AMR: American Ancestry n=347, EAS: East Asian ancestry n=503, EUR: European Ancestry n=502, SAS: South Asian Ancestry n=489). We conducted pairwise two-sample t-tests to assess whether the mean of the longer allele of each variant differed in size between each two populations. Only variants with a call rate above 80% in both of the two populations compared were taken into account. This resulted in the identification of a total of 438,512 population-specific variants (Bonferroni-adjusted P<4.4e-8) among at least one of the five superpopulations (*Supplementary Tab. 4*). As shown in *Supplementary Fig. 4A* and *Supplementary Tab. 5*, we detected 332,395, 226,226, 165,253, 119,282 and 59,654 STR variants which are specific to AFR, EAS, EUR, SAS, and AMR ancestry, respectively. Among these, 86,484 (19.7%) variants were unique to AFR, followed by 31,167 (7.11%) in EAS, 12,877 (2.94%) in EUR, 10,598 (2.41%) in SAS, and 5,020 (1.14%) in AMR ancestry. We also investigated the difference between the observed STR allele lengths in the 2504 unrelated samples and their corresponding loci on the reference genome (i.e., GRCh38). As expected, the empirical distribution of the observed data closely resembled a normal distribution (*Supplementary Fig. 4B*). As an illustrative example, we examined the distribution of the longer/shorter STR alleles at the *HTT* locus across 2,504 unrelated individuals from the 1000 Genomes Project, stratified by population (*Fig. 3C,D, Supplementary Fig. 3B,C*). This locus, located at chr4:3,074,876–3,074,939 on the GRCh38 reference genome, is known for its association with Huntington’s disease when expanded beyond a pathogenic threshold. As shown in *Fig. 3C*, the majority of individuals carry repeat lengths close to the reference allele (indicated by the red horizontal line), while population-specific deviations are evident. Notably, individuals of African ancestry (AFR) carried the largest number of repeats in the longer allele of *HTT*, whereas East Asian (EAS) populations exhibited comparatively fewer repeats in their longer alleles than other groups, reflecting underlying differences in repeat length variability at this locus. *Figure 3E,F* presents genome-wide Manhattan plots displaying the median SEI values for each STR variant across 3202 samples. Variants with higher median SEI values indicate loci with greater instability across individuals. Notably, chromosomes 2, 4, 10, 17, 20, 21, X, and Y harbor a higher density of unstable STR variants compared to other chromosomes, suggesting that these regions may be particularly prone to somatic expansion events.

**Figure 3.**
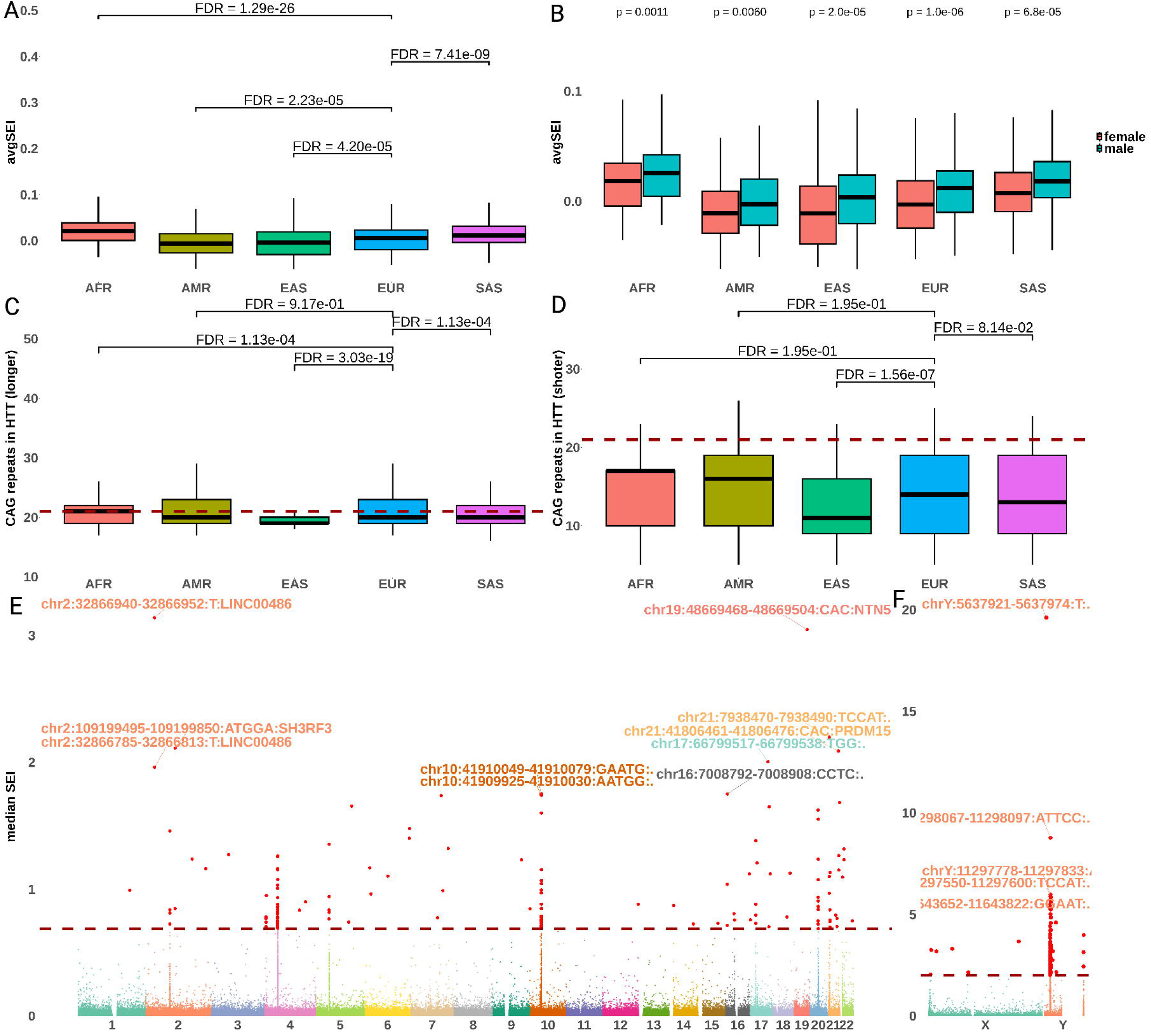
**(A)** Boxplot showing the genome-wide distribution of average SEI (avgSEI) stratified by five superpopulations from the 1000 Genomes Project: African ancestry (AFR), American ancestry (AMR), East Asian ancestry (EAS), European ancestry (EUR), and South Asian ancestry (SAS). (B) The distribution of avgSEI stratified by both the five superpopulation and sex. (C) Distribution of CAG repeats harbored in the HTT longer allele across the five superpopulations; red line indicates GRCh38 reference (chr4:3074876–3074939). (D) Distribution of CAG repeats harbored in the HTT shorter allele across the five superpopulations. Manhattan plots of median SEI of each locus across 3,202 1kGP samples in autosome, threshold=99.99th percentile (E), and sex chromosomes threshold= 99.9th percentile (F).

### Genotyping of short tandem repeats across 2,974 samples from the Rhineland Study

To evaluate the performance of *searchSTR* on target-enriched sequencing data, we applied the method to targeted sequencing data of 2,974 peripheral whole blood samples (male: 1,275, 43%, age range: 30-95, age mean ± standard deviation: 54.1±13.6) from the Rhineland Study. These samples were sequenced using Illumina NovaSeq 6000 with a targeted panel of 2,749 STR regions. A total of 4,823 candidate tandem repeat intervals were identified within the targeted region using *STRfinder*, which detects repeat motifs ranging from 1 to 10 bp in length. To ensure high-confidence genotyping, we filtered out redundant loci and those with low-complexity or ambiguous flanking sequences, resulting in a curated set of STRs suitable for short-read alignment-based genotyping. Afterwards, *searchSTR* was employed to genotype diploid STR alleles at each locus, leveraging its probabilistic framework to resolve diploid allele lengths from short-read sequencing data with high sensitivity. For benchmarking, we also applied *ExpansionHunter* to the same dataset and the corresponding repeat catalog. *SearchSTR* identified 2,994, 141, and 11 high-confidence STR variants on the autosomes, and the X- and Y-chromosome, respectively, with a per-locus call rate exceeding 75% (*Supplementary Tab. 6*). In comparison, *ExpansionHunter* identified 1677, 63, and 29 variants on the corresponding chromosomes under the same call rate threshold (*Supplementary Tab. 7*). As shown in *Fig. 4A* and *Supplementary Fig. 5*, *ExpansionHunter* frequently reported alleles that were substantially longer than their corresponding reference alleles and those estimated by *searchSTR*, including a notable number of extreme outliers (>100 repeat units). These inflated estimates were largely driven by poorly aligned reads, such as flanking reads with multiple mismatches in both the flanking and repeat regions. For example, at the *HTT* locus (chr4:3093905–3093929), REViewer visualizations revealed misaligned reads supporting artificially long alleles (*Supplementary Fig. 6*). This suggests a tendency toward overestimation of allele lengths at certain loci, particularly in the presence of repetitive or ambiguous sequence contexts. In addition, we assessed the SEI score for each STR locus in each individual based on its definition (*Online Methods, Equations 3,4*). A per-individual SEI score was then computed by averaging SEI values (avgSEI) across all loci, providing a genome-wide measure of somatic STR instability per sample (*Fig. 4B*).

**Figure 4.**
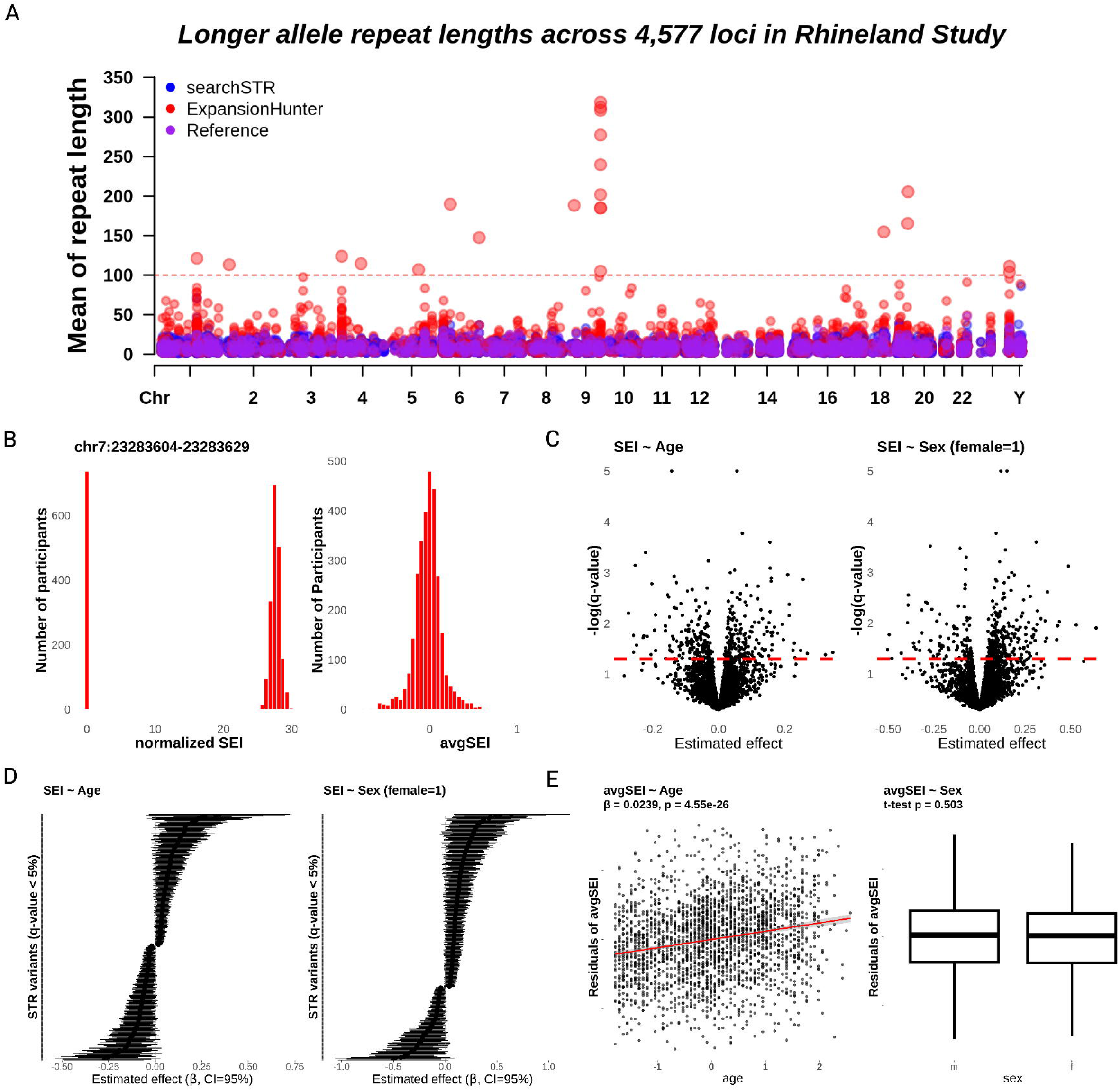
(A) Manhattan plots of mean repeat length of STRs in the longer allele in 2,974 participants of the Rhineland Study as estimated by searchSTR (blue), ExpansionHunter (red), and the corresponding reference genome allele sizes (purple), with outlier loci >100 repeat units highlighted. (B) Example of the distribution of log-normalized somatic expansion instability (SEI) scores at locus chr7:23283604–23283629, illustrating the zero-inflated normal distribution (left), and the distribution of average SEI (avgSEI) across individuals (right). (C) Volcano plots of STR SEI associations with age and sex, analyzed with zero-inflated Gaussian models, showing posterior mean regression coefficients versus –log₁₀(q-values) (sex coding: males=0, females=1); dashed lines mark significance at q = 0.05. (D) Forest plots of effect sizes and 95% credible intervals (CI) for STR variants whose SEIs were significantly associated with age or sex (q < 0.05) (sex coding: males=0, females=1); variant labels are provided in Supplementary Tab. 8. (E) Effects of age and sex on average SEI (avgSEI across 156 positively significant loci selected from Fig. 4D). No sex difference was observed.

### Age and sex effects on somatic expansion instability of STR variants

To investigate the influence of age and sex on SEI of tandem repeat variants, we modeled the association of SEI scores of 3,090 STR variants (autosomes: 2871, X-chromosome: 123, Y-chromosome: 96, each present in >50 individuals) with (endo)phenotypic variables of 2,964 individuals (10 out of 2974 had missing (endo)phenotype data) from the Rhineland Study, using a Bayesian zero-inflated Gaussian regression framework (*Online Methods*). First, in our models (*Equations 5-7*), we assessed the association of age and sex with SEI, reflecting their hypothesized roles in driving or modifying somatic STR expansion dynamics. To adjust for potential confounding due to ancestry-related population structure, we adjusted all models for the top 10 genetic principal components (PCs) derived from the paired genome-wide SNP data for each individual. As a result, SEI scores (*Equation 4*) for 179 STR variants showed a significant positive association with age, while 157 variants exhibited a significant negative association with age (q-value < 5%; *Fig. 4C*). For the analysis of sex effects, only the autosomal STR variants were considered to avoid confounding due to sex chromosome dosage differences. SEI scores for 227 STR variants were found to be significantly higher in females, while those of 96 variants were significantly lower at a q-value < 5%. The posterior effect sizes and corresponding 95% credible intervals for STR variants significantly associated with age and sex are presented in *Fig. 4D*. The full list of all STR loci whose SEI is statistically significantly associated with age and/or sex is provided in *Supplementary Tab. 8*. Furthermore, we analyzed the association between average SEI (avgSEI) and age using multiple linear regression, adjusting for sex and the top 10 genetic PCs as covariates. As shown in *Fig. 4E*, avgSEI, defined as the mean SEI across 156 of the 179 positively age-associated STRs (excluding loci on the sex chromosomes, *Supplementary Tab. 9*), exhibited a significant positive correlation with chronological age (β = 0.0239, p = 4.55 × 10⁻²⁶).

### Somatic instability effects on brain-related traits

To assess the potential contribution of STR somatic instability to brain aging and neurodegeneration, we examined the association of SEI with a set of brain-related traits, including plasma NfL levels and eight brain imaging-derived phenotypes (including total brain volume, total gray matter volume, supratentorial volume, hippocampal volume, subcortical gray matter volume, total cerebellar cortex volume, cerebral white matter volume, and mean cortical thickness). Specifically, we tested SEIs at each of 4,630 STR variants for association with these traits using 18 multivariable linear regression models, including two related to NfL and 16 related to brain imaging outcomes (*Online Methods and Supplementary Tab. 10*). All models were adjusted for potential confounders, including age, sex, the top 10 genetic PCs, and, specifically for volumetric outcomes, also estimated total intracranial volume (ETIV) to control for differences in head size. Models including NfL as the outcome were additionally adjusted for glomerular filtration rate to account for differences in kidney function, and in a sensitivity analysis, also for total brain volume. As shown in *Fig. 5* and *Supplementary Tab. 11-14*, we identified a set of 43 STR variants whose SEI showed statistically significant associations (FDR < 0.05) with at least one of the eight brain imaging-derived phenotypes (10 positive, 33 negative), and one STR variant (chr8:27568029-27568045:AAAAAATA (*GULOP*)) whose instability exhibited a significant positive association with plasma NfL levels in both models (*Supplementary Fig. 7*). In addition, we identified three other STR loci whose somatic instability interacted with age to either increase (chr19:37972168-37972206:GT (*SIPA1L3*)) or decrease (chr1:160483181-160483193:T and chr3:39060419-39060437:AT (*WDR48*)) plasma NfL levels. Among all 101 significant associations from the 18 models, 33 positive effects and 68 negative effects were directly associated with the SEI term, indicating highly locus-specific influences on brain-related traits (*Supplementary Tab. 13).* A set of novel STR variants shown in *Fig. 6A* may act as genetic modifiers of brain structure, particularly total gray matter volume and supratentorial volume. Notable examples include chr2:176094405–176094417:CA (*HOXD13*), chr5:95706837–95706849:AT (*CTD-2154I11.2*), chr19:47121845–47121857:CA (*SAE1*), and chr19:13490622–13490637:GAAAA (*CACNA1A*). Linear regression analyses between imaging phenotypes and avgSEI were conducted using 21 additional models (Supplementary Table 9). *Figure 6B* shows that higher avgSEI was significantly associated with reduced volumes of total gray matter, hippocampus, subcortical gray matter, and cerebellar cortex.

**Figure 5.**
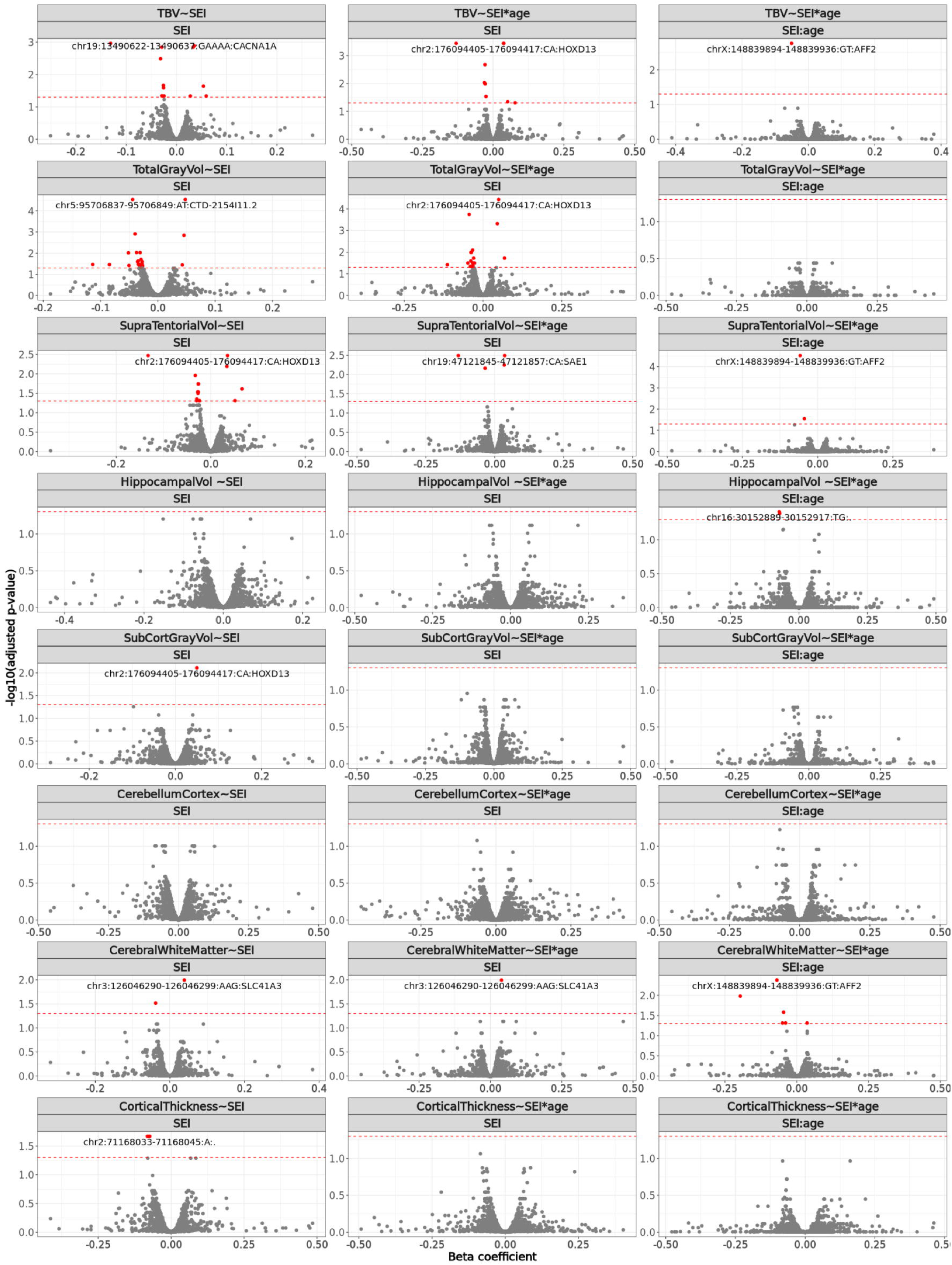
Volcano plots of the multiple regression-based estimates of the association between STR somatic expansion instability (SEI) (independent variable) and eight structural brain imaging-derived phenotypes (dependent variables). Each facet represents a separate regression model for a specific phenotype (e.g., TBV, cortical thickness, hippocampal volume), assessing the main effect of SEI or its interaction effect with age (SEI:age). The x-axis shows estimated effect sizes (beta coefficients), and the y-axis shows –log₁₀(FDR). Variants passing FDR < 0.05 (red dashed line) are marked in red, while the top variant is also labeled. Abbreviations: CerebellumCortex = total cerebellar cortex volume, CerebralWhiteMatter = cerebral white matter volume, CorticalThickness = mean cortical thickness, HippocampalVol = hippocampal volume, SubCortGrayVol = subcortical gray matter volume, SupraTentorialVol = supratentorial volume, TBV = total brain volume, and TotalGrayVol = total gray matter volume.

**Figure 6.**
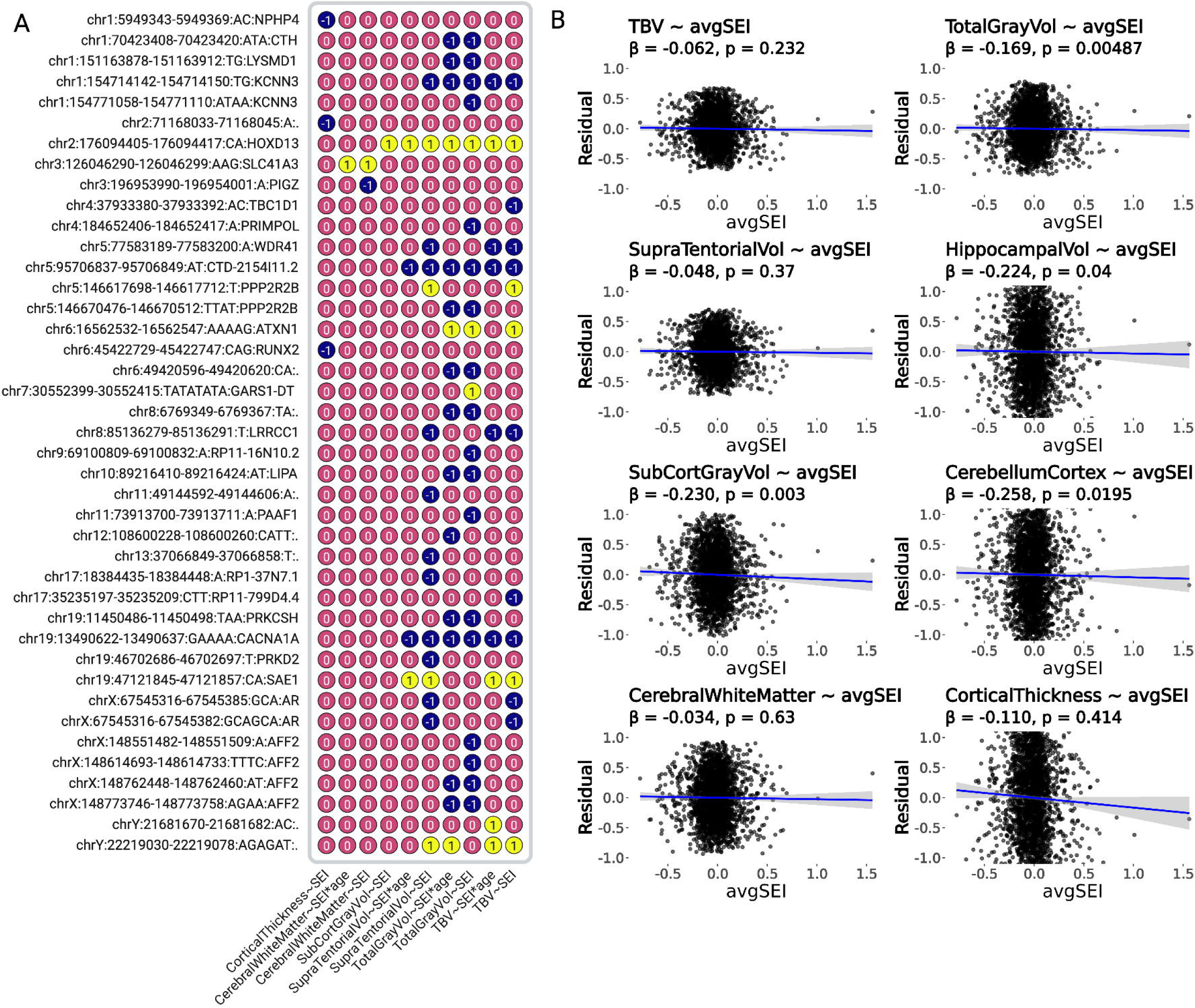
(A) Heatmap of STR variants whose somatic expansion instability (SEI) was statistically significantly associated with brain imaging-derived phenotypes. Colors indicate direction of association: yellow represents positive associations, and blue represents negative associations. Each cell corresponds to a variant–model pair, illustrating the pattern of effects across all models. (B) Partial effects of avgSEI (average SEI across the 156 selected loci) on eight brain imaging-derived phenotypes. The residuals represent the phenotype values after regressing out all other covariates except avgSEI. Abbreviations: 1 = positive association, −1 = negative association, Y∼SEI represents the effect of SEI of a variant on a phenotype Y, Y ∼ SEI * age represent the effect of the interaction between SEI and age on a phenotype Y. Abbreviations: CerebellumCortex = total cerebellar cortex volume, CerebralWhiteMatter = cerebral white matter volume, HippocampalVol = hippocampal volume, CorticalThickness = mean cortical thickness, SubCortGrayVol = subcortical gray matter volume, SupraTentorialVol = supratentorial volume, TBV = total brain volume, and TotalGrayVol = total gray matter volume.

## Discussion

Since the discovery of a CCG repeat expansion as the cause of fragile X syndrome in 1991, STR expansions have been found to cause over 60 other disorders, the majority of which are characterized by pronounced brain pathology [6]. Emerging evidence further indicates that age-associated somatic instability of these STR expansions represents a key mechanism in the pathogenesis of these disorders [9, 13, 44–46]. However, until now no computational methods existed for the systematic unbiased study of STR somatic instability. Here, we addressed this critical gap by developing an efficient, haplotype-resolved genotyping algorithm for high-throughput genome-wide profiling of somatic STR variations from either whole-genome or targeted sequencing data. Importantly, we have implemented this algorithm in the *searchSTR* tool, which is available through the software package *STRbean*.

Apart from an increased accuracy for calling STR genotypes, the most distinguishing feature of *searchSTR* compared to other existing tools is its ability to estimate STR somatic instability at genome-wide scale. This capability enabled us to systematically assess the association of SEI at many functionally relevant STR loci with a range of biologically highly relevant traits, including age, sex, and brain imaging-derived measures. To this end, we applied our approach to WGS data from 3,202 individuals of the 1000 Genomes Project as well as targeted deep-sequencing data from 2,974 participants of the Rhineland Study. This enabled us to generate a comprehensive multiancestry genome-wide reference panel of STR variants and their somatic instability, covering more than 1.4 million STRs across 26 populations. Remarkably, STR somatic instability at many loci was robustly associated with age, sex, brain morphology and markers of neurodegeneration.

Intriguingly, the associations of STR somatic instability with age were not uniform across the genome and highly locus-specific. Indeed, while we found that most STR loci did not exhibit somatic instability, SEI at many other loci robustly increased or decreased with age. Variants exhibiting higher SEI with age may reflect increased genomic instability at these loci in older individuals, potentially contributing to cellular dysfunction and neurodegenerative processes as recently found in Huntington’s disease [39, 40]. Conversely, negative associations with age may indicate either true age-associated decrease of SEI or reduced detectability of SEI at these loci in older individuals, possibly due to selective loss of cells carrying somatic expansions or age-related differences in DNA repair activity. For example, the expansion of an STR at chr9:136750347–136750359 (motif: AAAC), one of the loci showing the strongest age-related increase in instability, may disrupt the regulation of its nearest gene, *LCN6*. *LCN6* has been implicated in cerebrooculofacioskeletal syndrome 4, a nucleotide-excision repair disorder, and male fertility. These associations suggest that the accumulation of somatic STR expansions could contribute to such phenotypes. As the SEI at this locus increases with age, *LCN6* expression may progressively decrease, potentially leading to impaired DNA repair capacity, altered gene expression in neural and reproductive tissues, and reduced male fertility.

To the best of our knowledge, no previous reports exist on sex-specific differences in somatic STR instability. Similar to what we discovered for age, the association of sex with SEI was also highly locus-specific: There were more STR loci that exhibited a higher degree of somatic instability in females as compared to males. However, averaging SEIs across all autosomal loci revealed no significant sex difference. SEI values on the X and Y chromosomes exhibited higher densities than those on autosomes. In contrast to somatic instability, sex differences in germline instability of STRs are well established. For example, in Huntington’s disease, paternal transmissions are more often associated with further repeat expansions, whereas a strong maternal transmission bias has been found for congenital myotonic dystrophy type 1 [47, 48]. Similarly, a recent family-based study found distinct profiles of de novo mutations in STRs upon intergenerational transmission between males and females [49]. However, processes involved in spermatogenesis, ovogenesis or germ cell maintenance cannot account for somatic STR instability in blood-derived DNA. It therefore remains to be elucidated whether sex differences in DNA replication or repair processes may account for our findings.

Somatic instability at several STR loci was also associated with brain structure, including total brain volume, as well as volumes of gray and white matter and hippocampus as well as cortical thickness. The identified variants, many of which lie within or near genes previously implicated in neurodevelopment, synaptic function, or neuronal survival, point to potential mechanisms through which SEI may affect brain morphology. Notably, several of these genes have been linked to neurodevelopmental or neurodegenerative diseases, including Zimmermann-Laband syndrome (*KCNN3*), Ogden syndrome (*SAE1*), Joubert syndrome (*LRRCC1*), spinocerebellar ataxias (*ATXN1*, *CACNA1A* and *PPP2R2B*), fragile X syndrome (*AFF2*), and frontotemporal dementia/amyotrophic lateral sclerosis (*PFN1*). Significant interactions between SEI and age at some of these loci further indicated age-dependent effects. Interestingly, also here the effect directions were locus-specific and not uniform, suggesting that the influence of SEI on brain structure is strongly modulated by the STR’s genomic context. These findings reveal the importance of STR dynamics for the modification of age-related brain structure, and may partly explain inter-individual differences in susceptibility to neurodegeneration, laying the framework and furnishing the tools for future studies in this rapidly evolving area.

In our association studies, we restricted the analyses to spanning reads to estimate diploid repeat lengths as well as SEI, as they provide the most direct and reliable source of information for repeat size estimation. Nonetheless, our software also supports the incorporation of flanking and in-repeat reads, which can improve coverage and sensitivity at loci where spanning reads are limited. This functionality is particularly useful when sequencing read lengths are short (e.g., <100 bp) or when STR loci are very large, where spanning reads alone are often insufficient.

In conclusion, we present a novel computational framework and a suite of tailored bioinformatic tools for genotyping and characterizing STRs and their somatic instability using both whole-genome and targeted sequencing data. Our results underscore the importance of somatic STR instability in shaping the human brain. Moving forward, combining STR variation maps with other omics modalities—such as single-cell transcriptomes, chromatin accessibility, methylation, and proteomic data—may uncover the regulatory networks through which STR instability influences gene expression and neuronal integrity [50–53]. This integrative approach could open new avenues for understanding the molecular basis of brain aging and developing precision medicine strategies for neurodegenerative and psychiatric diseases.

## Data Availability

Whole genome sequencing CRAM files for 3202 1000GP samples, including 602 trios, 1598 males and 1604 females, aligned to GRCh38 were obtained from European Nucleotide Archive accessions PRJEB31736 (2504 original samples) and PRJEB36890 (698 additional related samples). Population and superpopulation labels for each sample were obtained from the 1000GP data portal (https://www.internationalgenome.org/dataportal/ sample). As described on the 1000GP data portal (https://www.internationalgenome.org/1000-genomes-summary/), all collections included in the 1000GP project followed their ethical guidelines and model informed consent language. The human reference genome GRCh38 can be accessed at https://console.cloud.google.com/storage/browser/_details/genomics-public-data/resou-rces/broad/hg38/v0/Homo_sapiens_assembly38.fasta. The data of the two Dutch cohorts used for the benchmarking analyses in this study have been reported previously [7]. The protocol of Rhineland Study was approved by the ethics committee of the University of Bonn Medical Faculty (Ref: 338/15). The study is conducted according to the International Conference on Harmonization Good Clinical Practice standards (ICH-GCP), with written informed consent obtained in accordance with the Declaration of Helsinki. The Rhineland Study's dataset is not publicly available because of data protection regulations. Access to data can be provided to scientists in accordance with the Rhineland Study's Data Use and Access Policy. Requests for further information or to access the Rhineland Study's dataset should be directed to. The genomic coordinates corresponding to the nine pathological polyglutamine STR loci used in the two Dutch cohorts were obtained from the Illumina Repeat Catalogs (available at: https://github.com/Illumina/RepeatCatalogs). STR catalogs are submitted to zenodo repository, and are accessible at [41, 42].

https://doi.org/10.5281/zenodo.11384920

https://doi.org/10.5281/zenodo.11383931

## Online Methods

### *STRfinder* algorithm

We developed an algorithm *STRfinder* (v1.0) that can exhaustively search for all possible STR patterns from a given sequence. To achieve this, *STRfinder* systematically scans the input sequence to identify STRs by searching for sequence patterns starting from the first base position and using a specified pattern size. Employing a sliding window approach, the algorithm iteratively shifts the window from left to right, quantifying the repeat length of the detected pattern. This process continues until no additional repeats are identified, allowing for a certain tolerance of base interruptions within the repeat sequence. This flexibility accommodates minor sequence variations or mutations, ensuring robust detection of STRs even in the presence of imperfect repeat patterns. If the identified pattern satisfies all filtering criteria, the corresponding locus is registered as an STR variant in a catalog, and the algorithm continues its search from the following base position after this STR locus. Otherwise, the algorithm resumes its search from the second base position, attempting to identify a new repeat pattern from the updated position. This process continues until the end of the sequence is reached. The same procedure is repeated for window sizes up to 6 bases, ensuring a comprehensive search for all possible STR loci. This iterative and exhaustive approach enables the accurate identification of STR variants across diverse sequence contexts. However, it may generate redundancy and overlapping intervals across different motif sizes. For instance, a variant containing 12 CAG repeats could also be identified as a variant with 6 CAGCAG repeats due to overlapping motif definitions. To address this issue, only variants with the smallest pattern size are retained in the catalog, ensuring that each STR is uniquely represented and eliminating redundancies caused by larger composite motifs. This approach prioritizes the most granular resolution of repeat patterns while maintaining consistency and clarity in the output. For overlapping intervals, a more refined strategy is applied: If the Jaccard index (i.e., the proportion of the overlapping region relative to the union of both intervals) is less than 50%, both intervals are retained. Conversely, if the Jaccard index exceeds 50%, only the longer interval is preserved. This method balances the need to capture distinct STR loci while minimizing redundant representation in cases of significant overlap. However, these overlapping intervals and closely adjacent STRs can lead to incorrect anchoring of variant boundaries during genotyping, as their flanking regions may contain a high repeat content. This issue becomes even more pronounced when the two adjacent STR patterns are similar. To resolve this issue, the algorithm annotates each STR with a sequence complexity score, using Shannon entropy and *k*-mer content, of its flanking sequences (within 10 bp and 20 bp) on both sides (see more in the section entitled *Filtering of STR loci for genotyping and downstream association studies*). STR variants with low-complexity flanking regions may be excluded using custom-designed criteria due to their high susceptibility to misalignment and false-positive read alignments. Additionally, each STR variant is annotated with the count of unknown bases (‘N’) present on both sides of the flanking regions within a distance of 1 kbp. To functionally characterize each locus, we provide the ability of annotating each STR based on GENCODE v38, when it falls within any region of any gene.

### Build catalogs of genome-wide STRs

We applied *STRfinder* (v1.0) to find and characterize STRs on the autosome and sex chromosomes of the human reference build GRCh38, generating two sets of STR catalogs (with ≤ 6 nucleotides in each repeat motif and ≥ 2 repeats): one ‘approximate’ catalog, and one ‘exact’ catalog. To construct the exact catalog, we configured *STRfinder* with the following parameters: -I hg38_reference _genome, -G GENCODE_v38, and -R 12 7 5 4 4 4. Here, -I specifies the input reference genome, - G provides the annotation file used for annotating identified STR loci, and -R defines the minimal repeat length for motifs of sizes 1 to 6 bp, respectively. For generating the approximate catalog, an additional parameter, -A, was included to enable approximate searches for repeat patterns.

### Collection of STR reads using *searchSTR*

To ensure comprehensive read collection for each STR locus, we systematically examined all mapped, mismapped, and unmapped reads that contain at least five consecutive repeat units. Following read selection, we applied a rigorous filtering process to identify high-confidence (“trustable”) reads, ensuring that only reliable sequences contribute to STR length estimation. Basically, our filtration process incorporates multiple genomic and sequencing features, including genomic coordinates, sequence quality, STR flanking regions, and mate-paired reads. For small STR alleles, our method can detect spanning reads that fully cover the repetitive sequences, enabling precise genotyping. However, for intermediate-sized STR alleles—where the repeat length exceeds individual read lengths but remains smaller than the overall fragment size—many collected reads are flanking reads, which only partially cover the STR allele. These flanking reads provide valuable information about STR expansions but require careful integration with spanning reads and paired-end data to accurately estimate the full repeat length. Moreover, we also collected in-repeat reads, which are fully contained within the STR allele, and have anchored mate reads in the nearby region. To account for genomic mutations and sequencing errors, repeat unit lengths are calculated with a predefined tolerance threshold, allowing for accurate inference of somatic STR expansions while minimizing false positives. To collect these reads in an efficient way for a set of targeted genome regions, we utilized the FM-index in SeqAn to provide an ultra-fast interface for searching a set of STR patterns within candidate reads. The FM-index, a compressed suffix array-based data structure, significantly enhanced the computational efficiency of our workflow, enabling scalable STR detection in large population cohorts. For genome-wide sequencing data, we used a hash map indexed by the aligned positions in the reference genome to efficiently identify candidate STR intervals for each well-aligned read.

### Inference of diploid genotypes using *searchSTR*

Although a set of trustable reads are collected for each STR locus as described above, these reads usually support more than two possible allele lengths in an individual, which is termed ‘scutter’ noise. The observed ‘scutter’ noise in STR genotyping can be attributed to multiple factors. One source of noise arises from DNA polymerase slippage at repeat loci during library preparation. However, a more biologically relevant source of variability is the age-related accumulation of somatic STR expansions. Over time, repeated DNA sequences become increasingly unstable, particularly in tissues with impaired DNA repair mechanisms. To provide a haplotype-resolved solution for STR genotypes, we applied a mixture of two skewed distributions to model spanning reads originating from the two STR alleles. Specifically, assume that the repeat lengths of the germline alleles are *µ* = *x*. Given a spanning read with *r* repeat patterns, the likelihood of *r* = *k* for this read will be *f*(*k*, *x*) = Pr(*r* = *k*|*µ* = *x*) ∼*p*(1 − *p*)^*d*^, where *d* = *µ* − *k* if *s* > *µ* − *k* ≥ 0, *d* = *β*(*k* − *µ*) if 0 > *µ* − *k* > −*s*, *d* = *s* if |*k* − *µ*| ≥ *s*. Here, *p* = 0.97 and *s* = 5 were empirically determined as the values that maximized Mendelian consistency of genotype calls in the Platinum Genome pedigree samples [54]. Unlike existing symmetrical models, we introduce a novel parameter *β* in this model, which can reflect the stability of somatic expansion. In this framework, a larger *β* indicates a larger degree of somatic instability. Existing genotype tools set *β* to 1 across all genome-wide loci. However, *β* in our model is a locus-specific parameter (*β* = 1.6 as the default). With the skewed model, the two-allele genotype then can be estimated by maximizing the likelihood of observing *r* = *k* within a mixture of the two skewed models with probability *π* belonging to the shorter allele: Pr(*r* = *k*|*µ*_1_ = *x*_1_, *µ*_2_ = *x*_2_) = *πf*(*k*, x_1_) + (1 − *π*)*f*(*k*, *x*_2_). For a flanking or in-repeat read containing *r* = *k* repeat units, the frequency function is given by *g*(*k*, *x*) = Pr(*r* = *k*|*µ* = *x*) ∼ 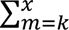 *f*(*m*, *x*), resulting in the likelihood of this observation within our mixture model of:

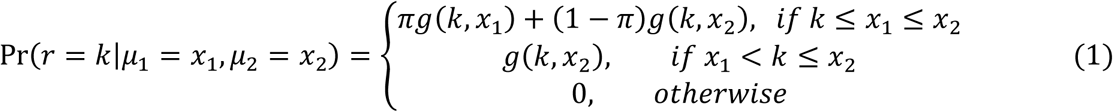

With all types of supporting STR reads, referred to as the set *S*, collected in the previous stage, the repeat lengths of the two alleles can then be estimated by maximizing the log-likelihood as follows:

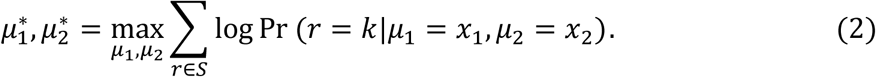

This optimization problem is computationally efficient to solve, as the repeat length is constrained to non-negative integers within a bounded range of *u*, where *u* represents the maximum observed number of repeat units. The bounded nature of the solution space significantly reduces the complexity of the optimization process, enabling efficient inference of haplotype-specific STR variants across a comprehensive, genome-wide catalog of STR loci. For STR alleles in Y- and X-chromosomes (in males), the number of repeats of the germline allele is taken to be the one with the highest frequency.

### Somatic expansion instability score for STR variants using *searchSTR*

To calculate individual-specific instability scores for each STR locus, the algorithm first estimates the repeat lengths of the two germline alleles as detailed in the section above. Subsequently, for a given STR locus *v* with a motif size of *m* bp, observed read counts of *x*_*vi*_ at each repeat length with i, j *ε* {1,2, ⋯ }, and the repeat length of the longer allele equal to *l*, the somatic expansion instability score (SEI), *S*(*v*), is defined as follows:

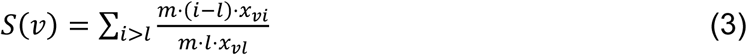

By standardizing the metric to the total number of reads harboring repeat sizes equal to *l*, we ensure that variations in the scores are primarily attributable to biological differences rather than technical or sampling biases, enabling robust comparative analyses across diverse individuals. Given the sparse nature of the SEI scores, which contain a large proportion of zero values, we applied an alternative log-transformation of the form,

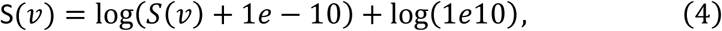

to normalize the distribution. This transformation preserves zero values (mapping 0 to 0), while effectively normalizing the non-zero SEI scores, resulting in a distribution more closely approximating normality for further downstream statistical analysis.

### Preprocessing and alignment of targeted sequencing data

Raw sequencing reads from the Rhineland Study participants were subjected to a standardized preprocessing pipeline to ensure data quality and consistency prior to STR genotyping. For each of the 2,974 samples, four FASTQ files (two sequencing lanes × paired-end reads) were generated. Initial quality control and adapter trimming were performed using TrimGalore [55](0.6.10) with default parameters, which wraps Cutadapt and FastQC to remove adapter sequences and low-quality bases from the reads. Post-trimming read quality was assessed using FastQC (v0.11.7), and all samples passed established quality thresholds for downstream analysis. High-quality trimmed reads were aligned to the human reference genome GRCh38 using BWA-MEM (v0.7.17-r1188), a Burrows-Wheeler aligner optimized for long and short paired-end reads. Alignments were processed with SAMtools (v1.20) to convert, sort, and merge BAM files across lanes for each sample. The resulting merged BAM files were then indexed using SAMtools, generating ready-to-use alignment files for downstream STR genotyping. This standardized preprocessing pipeline ensured high-quality and uniformly processed input data for comparative STR calling using both *searchSTR* and ExpansionHunter.

### Filtering of STR loci for genotyping and downstream association studies

Candidate STR loci were initially identified using *STRfinder*, which scans 2,749 customized targeted genome regions of the reference genome for tandem repeats with motif sizes ranging from 1 to 10 bp. During this process, *STRfinder* automatically removed redundant or overlapping repeat intervals to ensure non-redundant representation of STR loci. A total of 4,823 non-redundant loci were identified. To further improve the reliability of downstream genotyping, we implemented an additional filtering step based on the sequence complexity of flanking regions. For each candidate STR locus, we computed four sequence complexity scores – corresponding to the 10 bp and 20 bp regions immediately upstream and downstream of the repeat sequence. Loci exhibiting low-complexity flanking sequences on either side (as measured by Shannon entropy or sequence repetitiveness) were excluded from the final STR panel. This filtering step was designed to minimize alignment ambiguity and reduce genotyping errors due to poorly mappable or repetitive flanking regions, thereby enhancing the overall specificity and accuracy of STR calls from short-read data. Following genotyping, STR variants with less than 50 non-zero values in SEI scores were excluded to ensure data quality and reliability for downstream association analyses.

### Brain magnetic resonance imaging data of the Rhineland Study

Magnetic resonance imaging (MRI) scans were obtained using a 3T Siemens MAGNETOM Prisma system (Siemens Healthcare, Erlangen, Germany). The system is equipped with an 80 mT/m gradient and a 64-channel head-neck coil. T1-weighted images were acquired utilizing a multi echo Magnetization Prepared Rapid Acquisition Gradient Echo (MPRAGE) sequence with an isotropic spatial resolution of 0.8 mm. Post-processing was performed with our published protocol {Koch, 2025 #289} to derive brain structure volumes and cortical thickness.

### Association studies using linear regression models

To explore the association between somatic STR expansions and brain-related traits, we used multiple linear regression models, each evaluating the effect of SEI and avgSEI on a distinct phenotype. These phenotypes included circulating neurofilament light chain (NfL) levels and 8 brain structural measures derived from neuroimaging data, such as total brain volume (TBV), total gray matter volume (TotalGrayVol), supratentorial volume (SupraTentorialVol), average hippocampal volume (average_hip_vol), subcortical gray matter volume (SubCortGrayVol), total cerebellar cortex volume (total_cerebellum_cortex), cerebral white matter volume (cerebral_whitematter), and mean cortical thickness (mean_cortical_thickness), among others. Each of the 4,630 STR variants was tested independently for its association with each phenotype using data from 2,974 participants of the Rhineland Study. In each model, the phenotype was treated as the dependent variable, and the SEI score at a given STR locus served as the independent variable. All models were adjusted for relevant covariates, including age, sex and the top 10 genetic principal components (PCs, derived from individual-level genome-wide SNP data to account for population structure). The models with volumetric neuroimaging outcomes were also adjusted for estimated total intracranial volume (ETIV) to account for differences in head size, while the models with NfL as the outcome were additionally adjusted for blood volume and glomerular filtration rate, and as a sensitivity analysis, additionally for TBV to account for the amount of brain tissue from which NfL could originate. Additionally, we also included interaction terms between SEI and age (SEI × age) to assess whether the effect of somatic expansion on brain-related traits was modified by age. Continuous variables were z-standardized prior to regression modeling to ensure comparability of effect sizes across outcomes and loci. To ensure robustness of the regression models, influential outliers were identified and removed using Cook’s distance with a default threshold of Cook’s D > 4/n, where n is the sample size. We applied false discovery rate (FDR) correction to control for multiple testing across loci and traits. Statistical analyses were performed using the lm() function in R, and results were reported as standardized beta coefficients, standard errors, and adjusted p-values.

### Modeling age- and sex-associated patterns of somatic STR expansion using Bayesian zero-inflated Gaussian regression

A prominent characteristic of the SEI scores is the extremely high prevalence of zero values across individuals, reflecting loci with no indications for somatic instability. This sparsity of SEI (see in *Fig. 4B*) introduces specific challenges for conventional regression modeling. For example, standard linear or generalized linear models (GLMs) assume that the response variable follows a unimodal distribution (e.g., a Gaussian, Poisson, or negative binomial), and that variation in the response can be explained as a function of predictor variables such as age, sex, or STR locus features. However, when the majority of observations are structural zeros, these models tend to underestimate the frequency of zeros and misrepresent the underlying biological process. Moreover, the variance of the response is often overdispersed relative to the Poisson assumption, and may vary systematically with covariates. To address these challenges, we applied a zero-inflated Gaussian (ZIG) regression model for log-transformed SEI data (*Equation 4*). This framework assumes that each log-transformed SEI value is generated from a two-component mixture process: one component models the probability that a given observation is structurally zero (i.e., no detectable expansion), while the second models the distribution of continuous SEI scores when expansions are observed. In these models, we incorporated age and sex as the primary variables of interest. Age was modeled as a continuous variable, while sex was treated as a binary indicator. To control for population stratification and hidden structure in the genetic data, we additionally included the top 10 genetic PCs as covariates in the model. Formally, for individual *i* and STR locus *j*, let *y*_*ij*_ denote the (non-negative) log-transformed SEI score, yielding the following generative model:

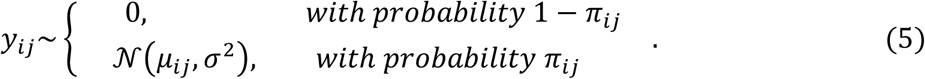

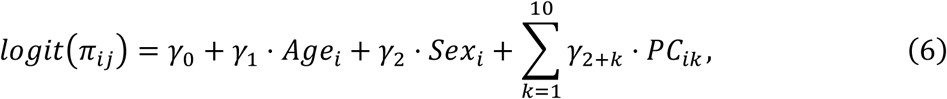

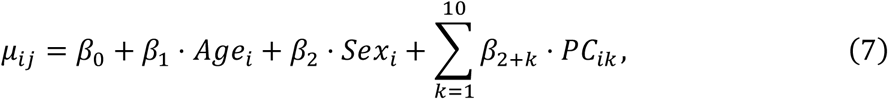

We placed informative priors on the regression coefficients to prevent overfitting while allowing flexibility: *β*_*k*_, *γ*_*k*_∼𝒩(0, 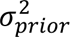) with *σ*_*prior*_ = 2 and 0.5, respectively. The residual variance *σ*^2^ of the Gaussian component is also assigned a prior *σ*^2^∼*lognormal*(0,1). We implemented the Bayesian zero-inflated Gaussian regression model using the brms package in R, which provides a high-level interface to Stan, allowing for efficient sampling of posterior distributions. Inference for model parameters was performed using Markov Chain Monte Carlo (MCMC) sampling that efficiently explores complex posterior distributions. Multiple chains (n=4) were run to ensure convergence. Posterior distributions of the regression coefficients for Gaussian components (*Equation 7*) were extracted to estimate the probabilistic effects of covariates such as age, sex, and the genetic PCs. To identify STR loci and covariate effects with robust statistical support, we applied Bayesian False Discovery Rate (Bayesian FDR) control based on the approach introduced by [56]. For each coefficient (e.g., age effect, sex effect) in the Gaussian model, we computed the posterior probability of a positive effect, defined as Pr(*β*_*k*_ > 0|*data*). Following Storey’s method, we treated these posterior probabilities as Bayesian analogues of local false discovery rates (lfdr) and derived q-values to control the expected proportion of false positives among discoveries. By calibrating across all tested loci, the Bayesian FDR procedure provides a principled and interpretable way to identify statistically credible associations between covariates (e.g., age, sex, PCs) and somatic expansion instability, while accounting for multiple testing across the genome. To determine the direction and significance of covariate effects on somatic STR expansion instability, we evaluated both the posterior mean of each regression coefficient and its corresponding q-value derived from Bayesian FDR control. Specifically, covariates with a posterior mean > 0 and q-value < 0.05 were classified as positively and significantly associated with increased SEI, indicating that higher values of the covariate are associated with greater somatic STR instability. Conversely, covariates with a posterior mean < 0 and q-value < 0.05 were classified as negatively and significantly associated with SEI, reflecting a protective or stabilizing effect. Covariates with q-values ≥ 0.05 were considered not statistically significant regardless of effect direction. Additionally, we analyzed the effect of age and sex on avgSEI using linear regression models (*Supplementary Tab. 9*).

### Tools used in this study

A variety of bioinformatics and statistical tools were employed throughout the study for preprocessing, STR detection, visualization, and Bayesian modeling. *ExpansionHunter* v5.0.0 was used to genotype known repeat expansions at targeted STR loci. FastQC {Andrews, 2010 #287} v0.11.7 was used for initial quality control assessment of raw sequencing reads. Cutadapt v5.1 was applied for adapter trimming and removal of low-quality bases from raw reads. BWA-MEM [57] v0.7.17-r1188 was used for aligning trimmed reads to the GRCh38 reference genome. SAMtools [58] v1.20 was used for sorting, indexing, and processing aligned BAM files. SeqAn [31] v3.0, a C++ template library for sequence analysis, was utilized as part of the *searchSTR* pipeline for detecting somatic STR expansions. Cmplot [59] v4.5.1 was used to generate Manhattan plots and chromosomal density plots to visualize genome-wide STR instability signals. brms v2.22.0, an R interface to Stan, was used to implement the Bayesian zero-inflated normal regression models for covariate effect inference using Markov Chain Monte Carlo (MCMC) sampling. REViewer [60] was used to visualize diploid estimations of STR alleles. All tools were run with default parameters unless otherwise specified. Analyses were conducted using a combination of in-house scripts in R (v4.3.2) and Python (v3.10), executed on a high-performance computing cluster.

## Code availability

Source code used in this study is available at https://github.com/jhu99/strbean and https://github.com/jhu99/SomaticInstability/.

## Acknowledgements

We would like to thank the Rhineland Study team for supporting the data acquisition and management as well as the PRECISE team for sequencing support. The Rhineland Study is funded by the German Center for Neurodegenerative Diseases (DZNE). Funding was also provided by the European Research Council (No. 101041677) and the National Natural Science Foundation of China (No. 62572398).

## Author contributions

JH, RM, MMB and NAA conceived and designed the project and the strategy. SE supported the collection and processing of the imaging data. KH, ED and MDB supported the generation of sequencing data. JH developed the computational package and performed the computation and statistical analysis. JH and NAA wrote the first draft of the manuscript. All authors reviewed and edited the manuscript.

## Supplementary information

Supplementary Figures 1-7

Supplementary Tables 1-14

---

**Supplementary Figure 1.** Benchmark analysis of STR length estimation for the shorter allele at nine polyglutamine disease–associated loci using searchSTR (cyan) and ExpansionHunter (red). The x-axis represents the ground truth (STR size measured by PCR-based fragment analysis). Each dot denotes a group of individuals with the same repeat size, with dot size indicating the number of samples in that group.

**Supplementary Figure 2.** (A) Detection rate of STR variants across the 1000 Genomes samples, with searchSTR identifying ≥1.2 million loci in most individuals. (B) Distribution of avgSEI values across autosomal loci, stratified by sex and superpopulation groups.

**Supplementary Figure 3.** Distribution of avgSEI (A), CAG repeats in the longer HTT allele (B), and CAG repeats in the shorter HTT allele (C) stratified by 26 populations. Red Line: number of HTT CAG repeats in the GRCh38 reference genome.

**Supplementary Figure 4.** (A) A Venn diagram illustrating population-specific and shared STR variants identified among 2,504 unrelated individuals from the 1000 Genomes Project (1kGP). (B) Distribution of STR difference in length between STR estimated by searchSTR and STR in reference.

**Supplementary Figure 5.** Mean STR lengths in the longer allele across the genome. Manhattan plots showing the mean repeat length per locus as determined by searchSTR (top panel), ExpansionHunter (middle panel), and the reference genome (bottom panel). Each point represents a single STR locus, positioned according to its genomic coordinates across chromosomes. Outlier loci with abnormally long alleles reported by ExpansionHunter are highlighted by a larger dot size.

**Supplementary Figure 6.** Read alignment visualization using REViewer of diploid estimations at the HTT locus (chr4:3093905–3093929:T) for one individual in whom ExpansionHunter suggested a pathological expansion. ExpansionHunter reported alleles exceeding 100 repeat units; however, the supporting reads exhibit poor alignment for the longer allele with mismatches in both the flanking and repeat regions, indicating likely overestimation.

**Supplementary Figure 7.** (A) Volcano plots of the multiple regression-based estimates of the association between STR somatic expansion instability (SEI) (independent variable) and neurofilament light chain (NfL) levels in plasma (dependent variable). Linear regression models were used to assess the main effect of SEI (left panel) or its interaction effect with age (SEI:age) (middle panel: main effect in the model including the interaction term, right panel: interaction effect). The x-axis shows estimated effect sizes (beta coefficients), and the y-axis shows – log₁₀(false discovery rate (FDR)-adjusted p-value). Variants passing FDR-adjusted p-value < 0.05 (red dashed line) are labeled. The x-axis shows estimated effect sizes (beta coefficients), and the y-axis shows –log₁₀(FDR). Variants passing FDR < 0.05 (red dashed line) are labeled. (B) Linear regression model to study partial effects of avgSEI on NfL in plasma.

