## Supplementary figures and images for "Genome-wide profiling of short tandem repeat somatic instability reveals associations with age, sex and brain-related traits"

### 7 - Supplementary_figure_1.png

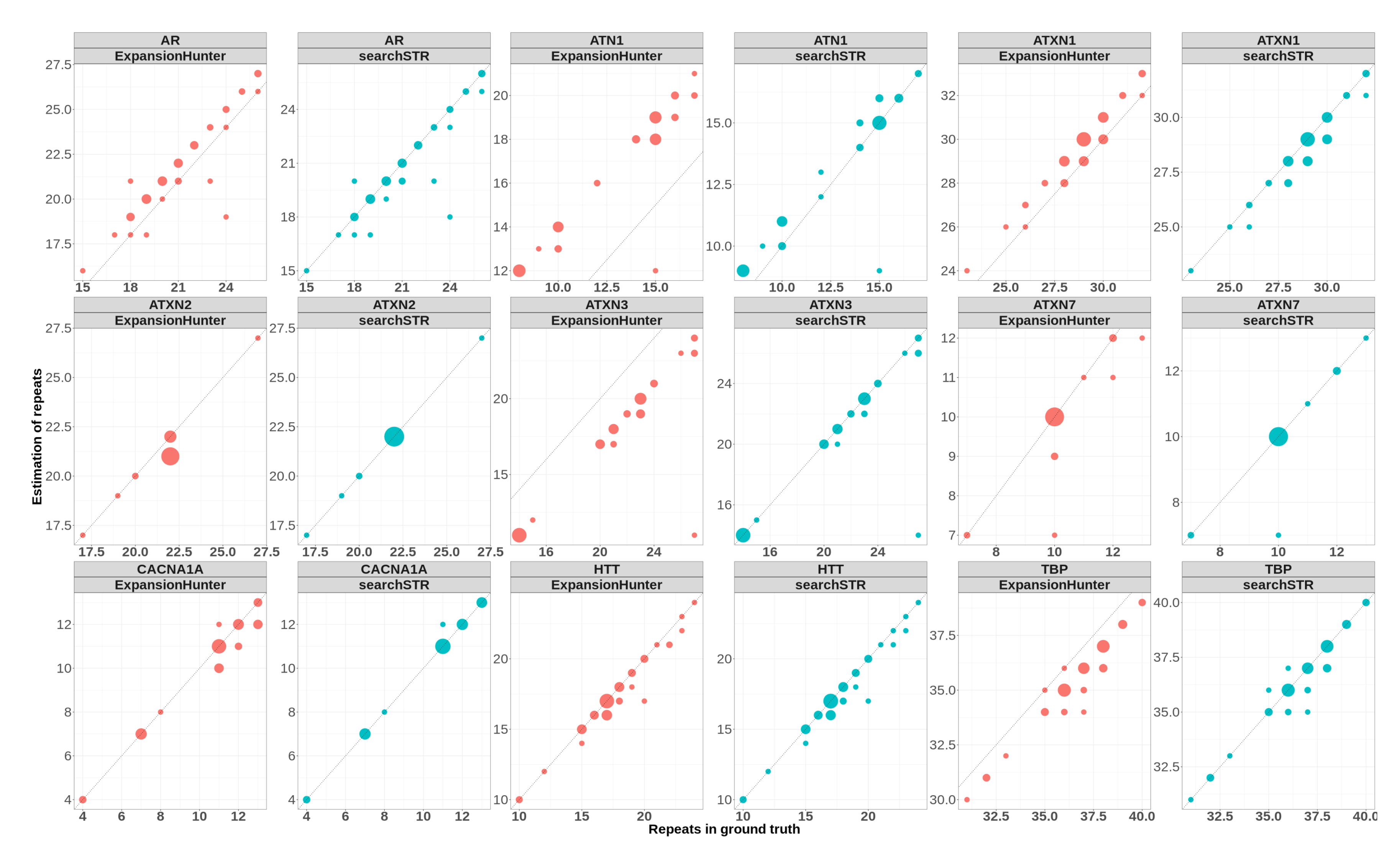

### 8 - Supplementary_figure_2.png

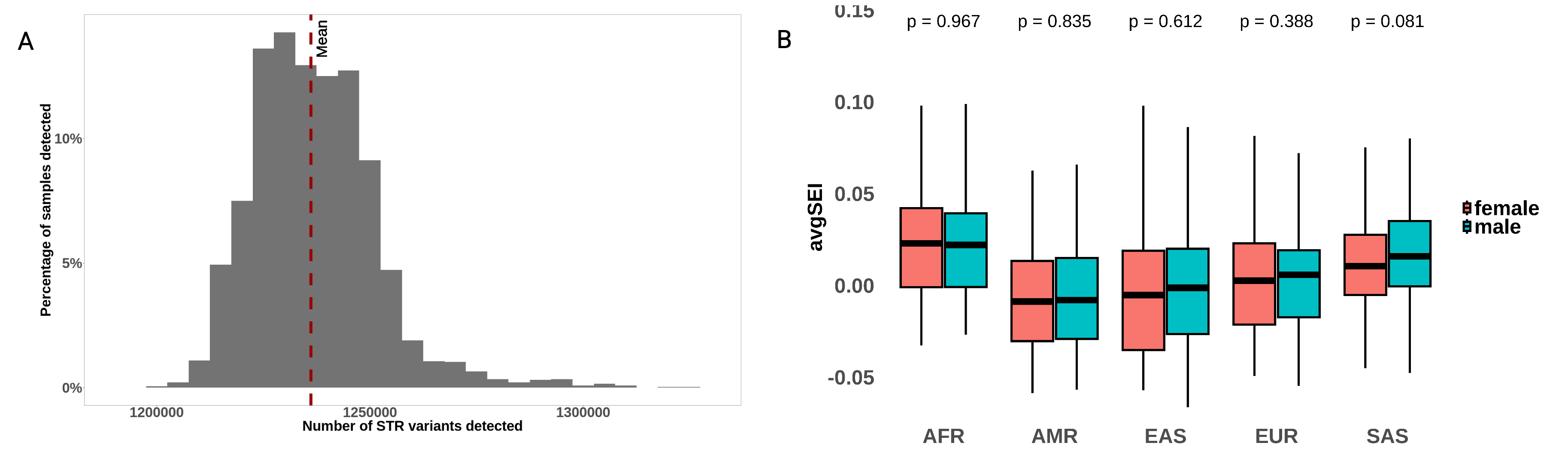

### 9 - Supplementary_figure_3.png

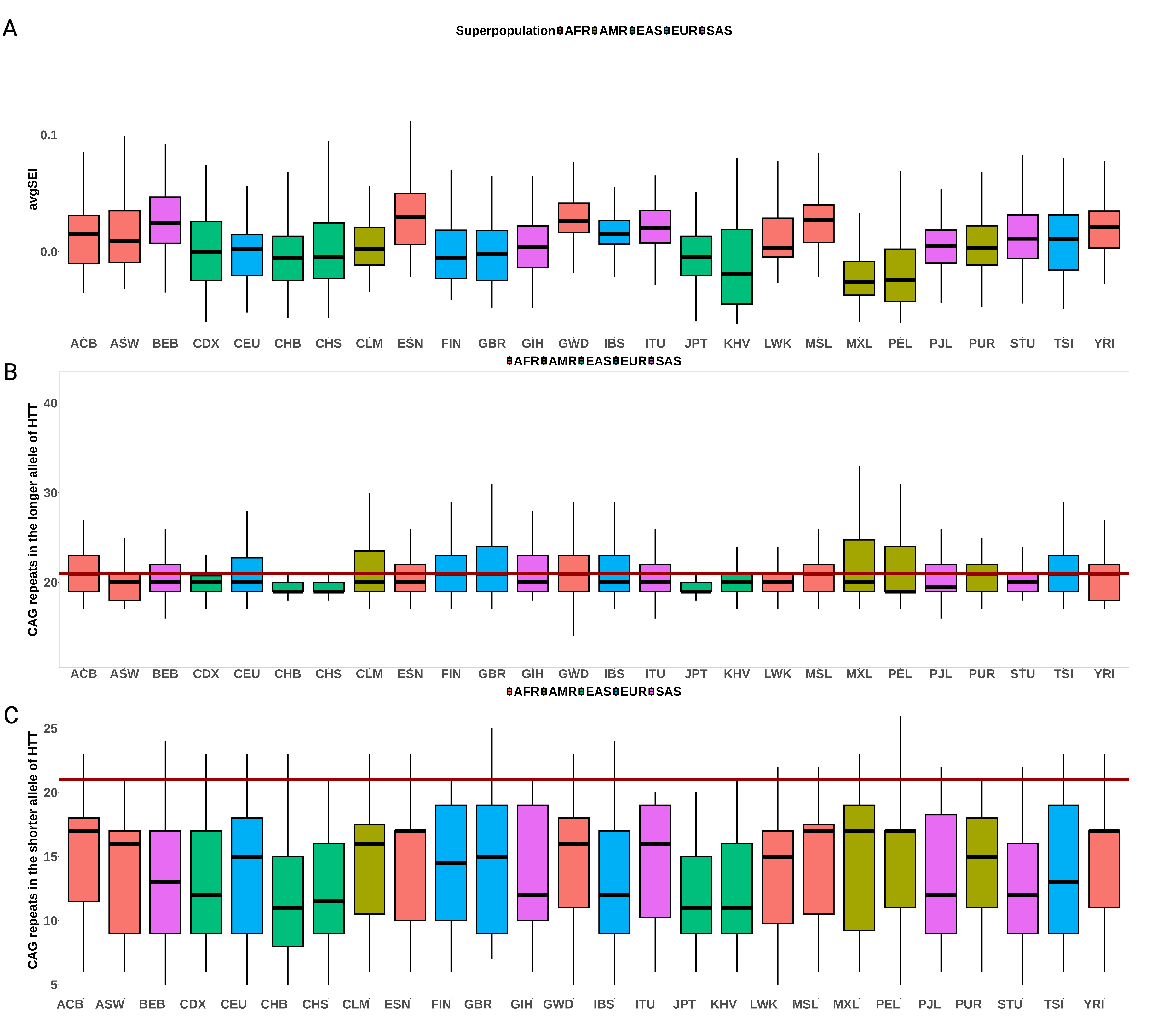

### 10 - Supplementary_figure_4.png

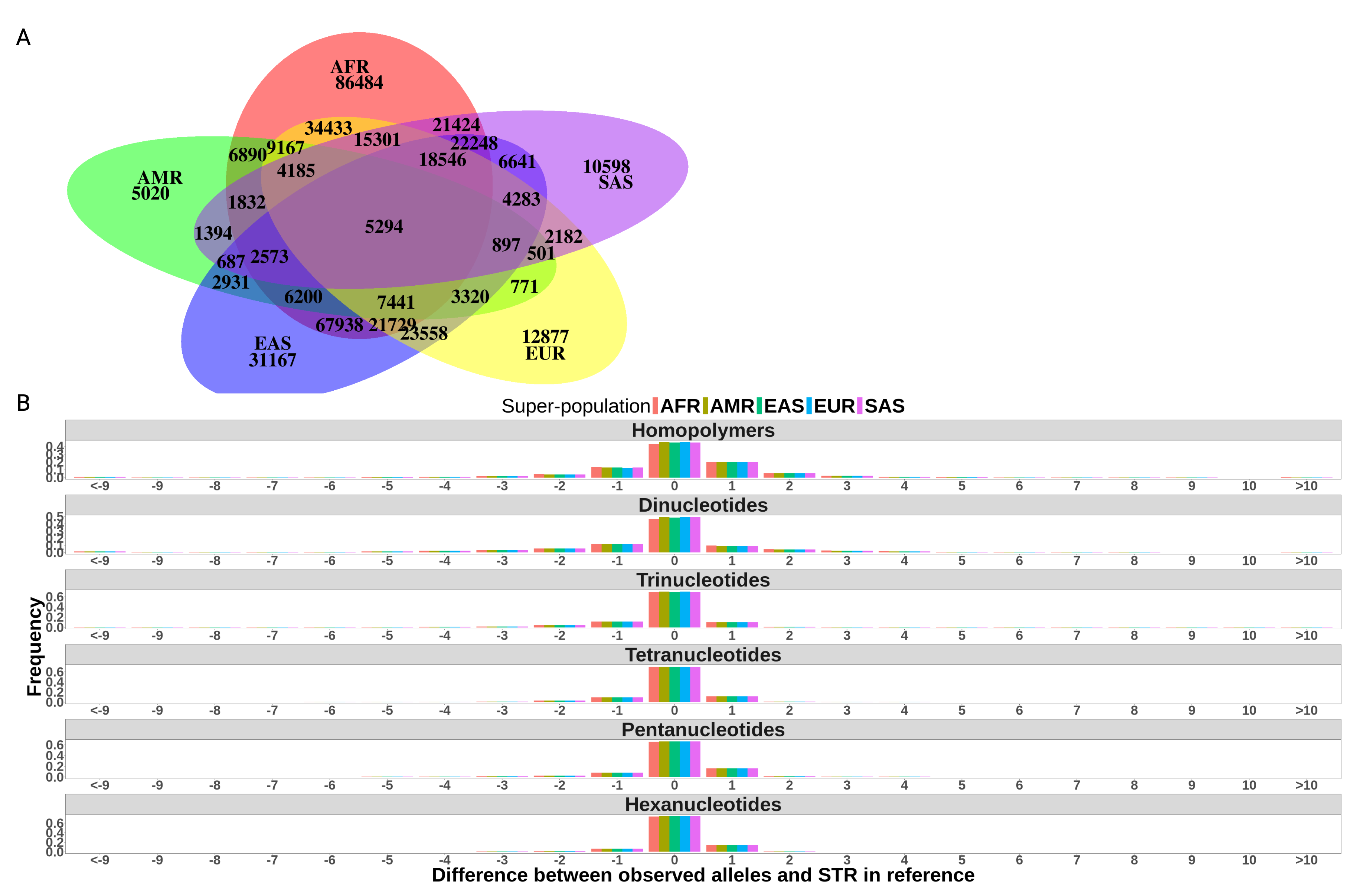

### 11 - Supplementary_figure_5.png

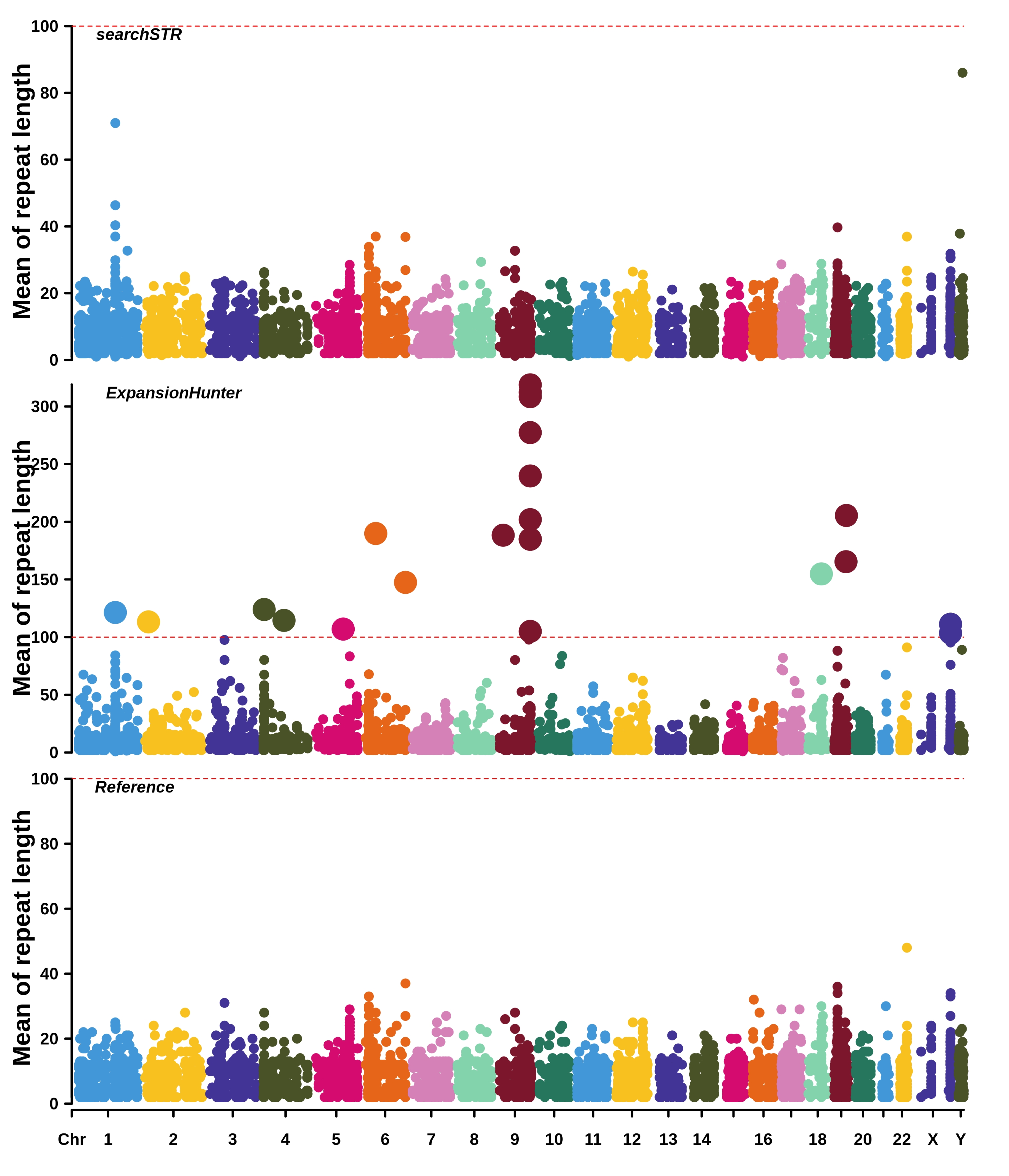

### 12 - Supplementary_figure_6.png

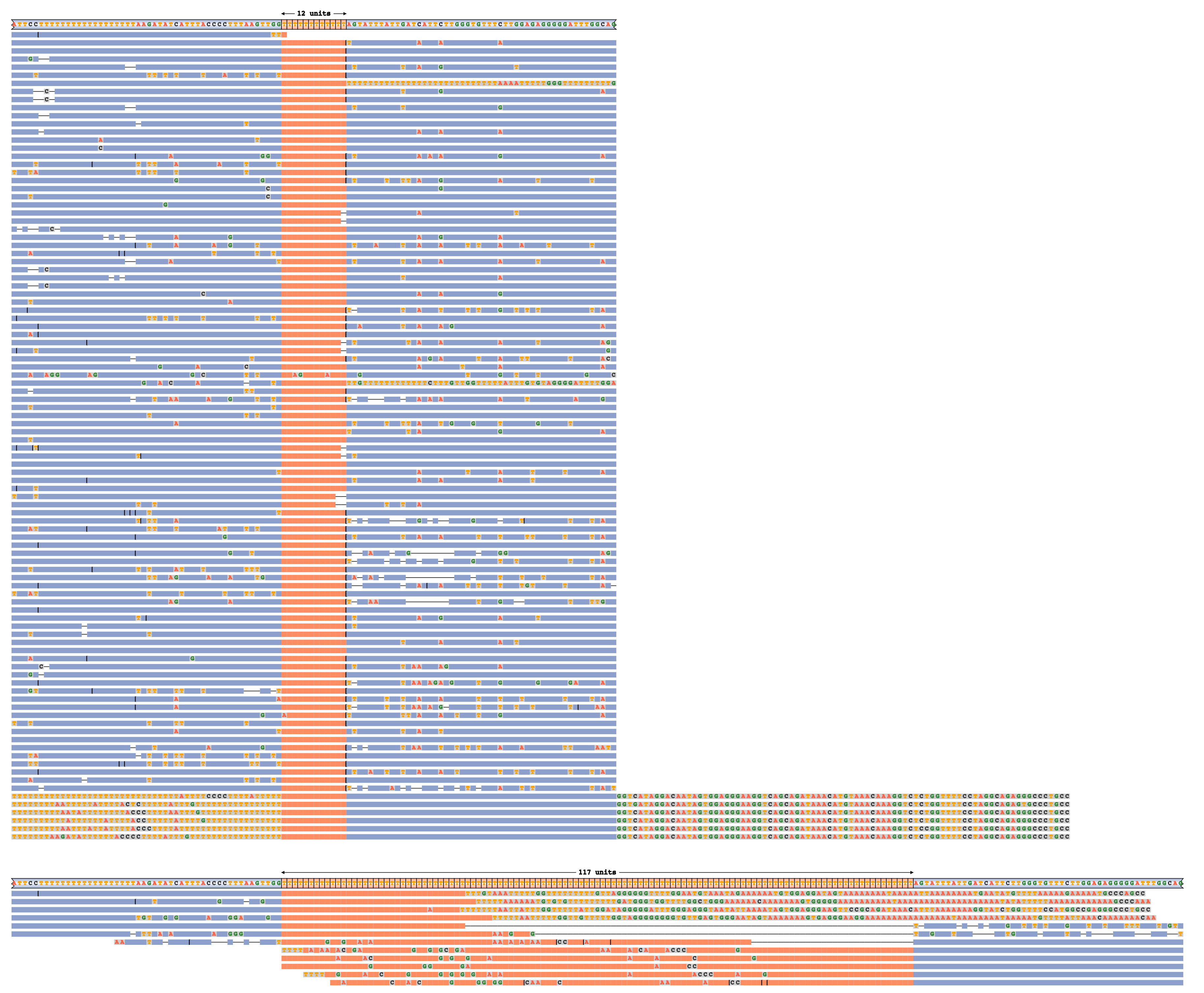

### 13 - Supplementary_figure_7.png

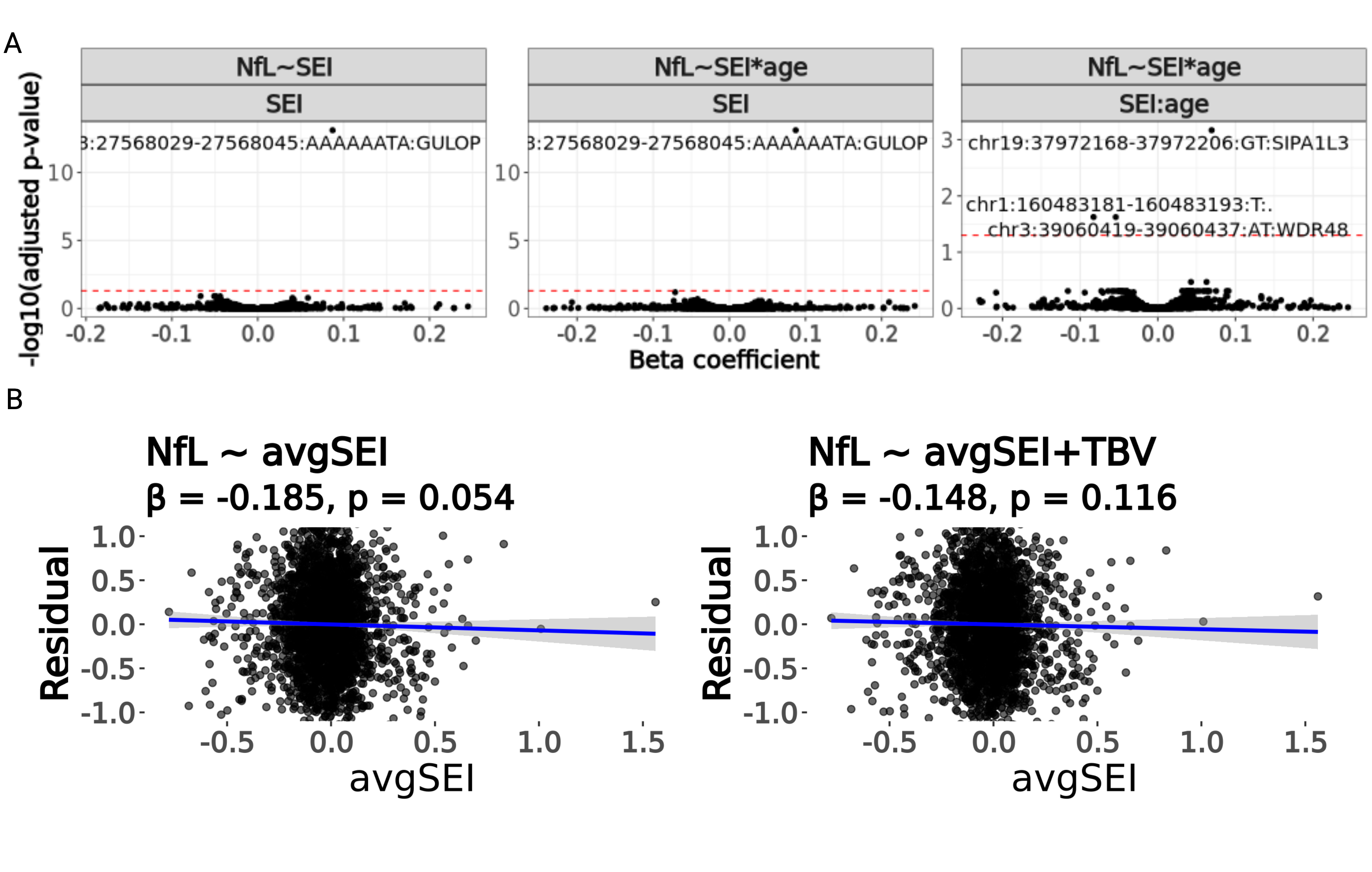
